# A genome-wide association study of adults with community-acquired pneumonia

**DOI:** 10.1101/2023.08.29.23294759

**Authors:** Eva Suarez-Pajes, Itahisa Marcelino-Rodriguez, Elisa Hernández Brito, Silvia Gonzalez-Barbuzano, Melody Ramirez-Falcon, Eva Tosco-Herrera, Luis A. Rubio-Rodríguez, María Luisa Briones, Olga Rajas, Luis Borderías, Jose Ferreres, Antoni Payeras, Leonardo Lorente, Javier Aspa, Jose M. Lorenzo Salazar, José Manuel Valencia-Gallardo, Edurne Carbonell, Jordi Freixinet, Felipe Rodríguez de Castro, Jordi Solé Violán, Carlos Flores, Carlos Rodríguez-Gallego

**Affiliations:** Research Unit, Hospital Universitario Nuestra Señora de Candelaria, Santa Cruz de Tenerife, Spain; Area of Preventive Medicine and Public Health, Universidad de La Laguna, Santa Cruz de Tenerife, Spain; Department of Immunology, Hospital Universitario de Gran Canaria Dr. Negrín, Las Palmas de Gran Canaria, Spain; Genomics Division, Instituto Tecnológico y de Energías Renovables (ITER), Santa Cruz de Tenerife, Spain; Department of Respiratory Diseases, Hospital Clínico y Universitario de Valencia, Valencia, Spain; Department of Respiratory Diseases, Hospital Universitario de la Princesa, Madrid, Spain; Department of Respiratory Diseases, Hospital San Jorge, Huesca, Spain; Critical Care Unit, Hospital Clínico de Valencia, Valencia, Spain; INCLIVA Biomedical Research Institute, Valencia, Spain; Department of Internal Medicine, Hospital Son Llatzer, Palma de Mallorca, Spain; Intensive Care Unit, Hospital Universitario de Canarias, San Cristóbal de La Laguna, Spain; Department of Respiratory Diseases, Hospital Universitario de Gran Canaria Dr. Negrín, Las Palmas de Gran Canaria, Spain; Department of Thoracic Surgery, Hospital Universitario de Gran Canaria Dr. Negrín, Las Palmas de Gran Canaria, Spain; Department of Medical and Surgical Sciences, School of Medicine, Universidad de Las Palmas de Gran Canaria, Las Palmas de Gran Canaria, Spain; Department of Clinical Sciences, University Fernando Pessoa Canarias, Las Palmas de Gran Canaria, Spain; Critical Care Unit, Hospital Universitario de Gran Canaria Dr. Negrín, Las Palmas de Gran Canaria, Spain; CIBER de Enfermedades Respiratorias (CIBERES), Instituto de Salud Carlos III, Madrid, Spain

## Abstract

**Introduction:** Community-acquired pneumonia (CAP) is associated with high morbidity and hospitalization rate. In infectious diseases, host genetics plays a critical role in susceptibility and immune response, and the immune pathways involved are highly dependent on the microorganism and its route of entry.

**Objective:** To identify genetic risk loci for CAP using a one-stage genome-wide association study (GWAS).

**Methods:** We performed a GWAS on 3,765 Spanish individuals, including 257 adult patients hospitalized with CAP and 3,508 population controls. Pneumococcal CAP was documented in 30% of patients; the remaining 70% were selected among patients with unidentified microbiological etiology. Genotyping and imputation allowed testing the association of 7,6 million variants using logistic regressions. Subsequently, we prioritized genes and likely causal variants based on Bayesian fine mapping and functional evidence. Imputation and association testing of the classic HLA alleles and amino acids was also conducted.

**Results:** We revealed six independent sentinel variants that were genome-wide significant (*p*<5×10^−8^), three located on chromosome 6p21.32, and one for each of the chromosomes 4q28.2, 11p12, and 20q11.22. Our analyses prioritized *C4orf33* on 4q28.2, *TAPBP* on 6p21.32, and *ZNF341* on 20q11.22. Interestingly, genetic defects of *TAPBP* and *ZNF341* are previously known inborn errors of immunity predisposing to bacterial pneumonia, including pneumococcus and *Haemophyilus influenzae*. Associations were all non-significant for the classic HLA alleles.

**Conclusions:** We completed a GWAS of CAP and identified four novel risk loci involved in CAP susceptibility.

## INTRODUCTION

Infections are one of the main causes of death globally and nearly one in eight deaths continue to be due to bacterial and viral infections (Lanks et al., 2019). Community acquired pneumonia (CAP) is considered a major public health problem due to its high morbidity and mortality (Mortensen & Metersky, 2012; Theilacker et al., 2021). Yearly CAP incidence varies widely worldwide, in Europe it ranges from 20.6/10,000 in Iceland (Bjarnason et al., 2018) to 79.9/10,000 persons in the UK (Millett et al., 2013). The available data for adults in Spain estimates 46.3 cases per 10,000 inhabitants (Rivero-Calle et al., 2016). One-ninth of hospitalized patients with CAP require admission to the intensive care unit (ICU) due to complications such as severe respiratory failure or septic shock (Ranzani et al., 2017; Rello & Perez, 2016). In fact, mortality is restricted mostly to the patients that are hospitalized (6–20%) (Ramirez et al., 2017; Welte et al., 2012), which could be as high as 50% among those admitted to the ICUs (Rello & Perez, 2016).

Host genetics plays a central role in the response against pathogens and contribute to explain the differences in susceptibility and severity among patients (Casanova & Abel, 2022; Ishak et al., 2022; Lees et al., 2019). Specifically, SNP-based heritability assessments in pneumonia support that genetic host factors explain a greater proportion of severity than of susceptibility (Chen et al., 2021). However, there is a paucity of genetic studies aimed at identifying genetic factors involved in CAP susceptibility or prognosis. In addition, most of them have focused on candidate genes, especially on genes involved in the immune response such as those encoding the mannose-binding lectin (*MBL*), the surfactant proteins A (*SFTPA1*, *SFTPA2*) and D (*SFTPD*), or the interleukins (*IL-6*, *IL-10*), to name a few (Gallagher et al., 2003; Garcia-Laorden et al., 2008; García-Laorden et al., 2011; Kloek et al., 2019; Schaaf et al., 2005; Solé-Violán et al., 2011). A few other studies have relied on genome-wide association studies (GWAS) to reveal pneumonia susceptibility loci in the HLA, *MUC5AC*, *IL6R*, and *TNFRSF1A* (Reay et al., 2022; Tian et al., 2017), and pneumonia severity loci in *CFTR*, *R3HCC1L*, and *HBB* (Chen et al., 2021). However, it must be noted that typically these studies have not distinguished the source of infection, implying that the patients are a heterogeneous mixture of patients with CAP and with nosocomial infections, i.e., with hospital-acquired pneumonia (HAP).

CAP is typically caused by several bacteria, including *Streptococcus pneumoniae* and *Haemophylus influenzae,* or viruses (Gadsby et al., 2016; Lanks et al., 2019; Musher et al., 2017; Su et al., 2023). *S. pneumoniae* is the leading cause of CAP and causes about one-third of hospitalized cases in Europe, although these frequencies are underestimated (Dion & Ashurst, 2022; Gadsby et al., 2016; Johansson et al., 2010; Musher et al., 2017; Pick et al., 2020; Torres et al., 2018, 2021). The immune response varies widely depending on the causative pathogen. Inborn errors of immunity (IEI, usually referred to as primary immunodeficiencies) strongly support that predisposition to infection by different microorganisms usually relies on different components of the immune system. For example, IEI impairing type I interferon-mediated immunity predispose to susceptibility to severe pneumonia by SARS-CoV-2 or influenza viruses, whereas IEI predisposing to pneumococcal infection are particularly involved in opsonization or phagocytosis of opsonized bacteria by splenic macrophages (Boisson, 2020; Casanova & Abel, 2021, 2022; Picard et al., 2003; Su et al., 2023). Therefore, studies aimed to identify the genetic basis of susceptibility or severity of infections may benefit from a precise homogenization of the source of infection and the causative microorganism.

Here we have conducted a GWAS of hospitalized patients with the only diagnosis of CAP, focusing on patients with pneumococcal infection or without identified causal microorganism.

## MATERIAL AND METHODS

### Study design

We conducted a one stage case-control GWAS of adult subjects of European ancestry from Spain. A total of 259 adult hospitalized patients with CAP diagnosis were recruited between March 2001 and 2016 from six Spanish hospitals and constituted the cases. It is assumed that pneumococci cause most CAP cases in which negative test results were found using conventional microbiological methods (Gadsby et al., 2016; Johansson et al., 2010; Torres et al., 2018). Therefore, to keep homogeneous the causative microorganism of CAP, only patients with confirmed pneumococcal infection or those in whom no identified causative microorganism was identified were included. These cases have been used in previous candidate gene association studies of CAP (Garcia-Laorden et al., 2008; García-Laorden et al., 2011, 2020; Solé-Violán et al., 2011, 2021). The study inclusion criteria and phenotype descriptions are available in the **Supplementary material**. The DNA from cases were extracted from whole blood samples combining standard phenol-chloroform procedures with automated column-based nucleic acid purifications as previously described (Garcia-Laorden et al., 2008; García-Laorden et al., 2011, 2020; Solé-Violán et al., 2011, 2021).

As controls, we used the genetic data available from 3,526 donors from the Spanish DNA Biobank (https://www.bancoadn.org), collected from the National Blood Service and have been used in a previous GWAS of severe COVID-19 (Cruz et al., 2022). All control participants were clinically uncharacterized unrelated adults that self-reported being of Spanish origin and the absence of personal or familial history of diseases, such as infectious, cancerous, circulatory, endocrine, mental, or behavioral, nervous, visual, auditory, respiratory, and immunological, among others.

Written informed consent was obtained from all participants or their representatives. The study was conducted according to The Code of Ethics of the World Medical Association (Declaration of Helsinki), and the Research Ethics Committees from the coordinating centers approved this study (Hospital Universitario de Gran Canaria Dr. Negrín Ethics Committee FIS PI 16/00759; Hospital Universitario Nuestra Señora de Candelaria Ethics Committee PI-19/12).

### Genotyping, quality control, and variant imputation

Both cases and controls were genotyped with the Axiom Spain Biobank Array (Thermo Fisher Scientific) following the manufacturer’s instructions in the Santiago de Compostela Node of the National Genotyping Center (CeGen-ISCIII; http://www.usc.es/cegen). This array contains a total of 757,836 variants among which rare variants selected from the Spanish population are included. Variant calling was performed using the Axiom Analysis Suite software following the manufacturer’s recommendations.

The quality control steps were conducted for the cases and controls considered together using PLINK v.1.9 (Chang et al., 2015) and R v.4.1.1 (R Core Team, 2022) for the variants located in autosomes. The filtering steps excluded the genetic variants with a minor allele frequency (MAF) <1%, a genotyping rate <95%, or showing evidence of large Hardy-Weinberg equilibrium deviations (HWE, *p*<1.0×10^−6^). Moreover, the individuals with some degree of relatedness (PIHAT > 0.2), deviations in heterozygosity, or incomplete clinical data, were excluded from the study. We also conducted a principal component (PC) analysis (PCA) with PLINK to discard genetic outliers and to derive the main PCs to then correct for the effect of the population stratification in the stage of association modelling. These procedures left us with a total of 603,603 genetic variants for 257 hospitalized CAP patients and 3,508 controls.

Variant imputation was conducted on the resulting dataset using the Michigan Imputation Server, relying on Eagle v.2.4 (Loh et al., 2016) for pashing and the Haplotype Reference Consortium release 1.1 (2016) as the reference panel (McCarthy et al., 2016).

### Statistical analysis and the functional assessment of associated loci

#### Variant association testing

To test the association of genetic variants with CAP, we used additive logistic regression models across all the imputed variants satisfying a good imputation quality (Rsq≥0.3) and a MAF≥1% using EPACTS v.3.2.6 (http://genome.sph.umich.edu/wiki/EPACTS). The association model was adjusted for sex and the first 3 PCs and the results were assessed using the genomic inflation factor (λ) calculated with the *gap* package for R. Variant associations were considered statistically significant at a threshold *p*<5.0×10^−8^. Independent sentinel variants were identified as those surpassing *p*<5.0×10^−8^ and showing weak linkage disequilibrium (r <0.1) with others in each locus after clumping in PLINK.

A sensitivity analysis of the sentinel variants was conducted by including other covariates in the logistic regression model (e.g., age), and also testing their association with other two recorded severe pneumonia outcomes in the case series by separate and combined: i) severe sepsis or septic shock according to the criteria available at the moment of the start of the recruitment (Bone et al., 1992); and ii) severe respiratory failure, defined as oxygen saturation <90% on room air, or a partial pressure of oxygen [PaO_2_]<60 mmHg.

#### Bayesian fine mapping

We performed a fine mapping on the association results around the independent sentinel variants to identify the credible variant set that most likely harbors the causal variant with 95% confidence, assuming that there is only one causal variant in each locus and that such variant has been tested in the study. For this, we used the *corrcoverage* package (Hutchinson et al., 2020) for R to calculate the posterior probabilities of the variant being causal for all polymorphisms within a region of 2 Megabase pairs with the leading variant and had an r >0.1 with it. The variants were considered part of the credible set until their sum of probabilities was ≥ 0.95.

#### Functional analysis of the variants

To assess the biological consequences of the variants included in the credible sets of the associated loci, we functionally annotated the variants using Ensembl Variant Effect Predictor (VEP) v.105 to obtain the scaled Combined Annotation Dependent Depletion (CADD) score v.1.6 of each variant. In addition, we used the Variant-to-Gene (V2G) score to prioritize the genes that were most likely affected by the functional evidence based on expression quantitative trait loci (eQTL), chromatin interactions, *in silico* functional predictions, and distance between the prioritized variants and transcription start site (TSS), based on data from the Open Targets Genetics portal (Ghoussaini et al., 2021). To predict the biological impact of the variants on the prioritized genes, we used the Mutation Significance Cutoff (MSC) v.1.6 of genes (Itan et al., 2016) with a confidence interval of 99% to interpret the CADD score. For those variants deemed to predict a high biological impact (with a CADD > MSC), we performed an *in silico* analysis to determine their potential regulatory roles, including the effects in enhancer and promoter histone marks, DNase I accessible sites, and other altered motifs by using HaploReg v.4.2 (Ward & Kellis, 2012) and RegulomeDB (Boyle et al., 2012).

To assess tissue expression of the genes prioritized in the associated loci and the existence of expression quantitative trait loci (eQTL) in the sentinel variants on artery, esophagus, lung, and whole blood, we used The Genotype-Tissue Expression (GTEx) Release v.8 data (https://www.gtexportal.org/home/). In parallel, we accessed a public transcriptomic dataset (GSE65682) available from the Gene Expression Omnibus (GEO) to assess expression differences in the genes prioritized in the associated loci among 108 ICU patients with CAP diagnosis and 42 healthy controls as described in the study by Scicluna et al (Scicluna et al., 2015). The statistical differences in gene expression between healthy controls and the patients with sepsis by CAP were evaluated using a *t*-test for the prioritized genes on each locus. Detailed information of this analysis is available in the **Supplementary material**.

### Association of classic HLA alleles and amino acids

Due to the sequence complexity of the human leukocyte antigen (HLA) complex, its key role in immunity, and the previously reported association with pneumonia susceptibility (Tian et al., 2017), we also performed a targeted association testing of the variation in genes of the HLA complex with CAP. To this end, we imputed the genetic variation at eight classical HLA genes (−*A*, −*B*, −*C*, −DPA1, −*DPB1*, −*DQA1*, −*DQB1*, and −*DRB1*) with the Michigan Imputation Server using four-digit Multi-ethnic HLA v.1 as a reference panel (Luo et al., 2021). Subsequently, we tested the association of the amino acids and the classical four-digit HLA alleles for the alleles with a frequency >1%. For this purpose, we followed the workflow described in HLA-Tapas (Luo et al., 2021), and the significance was established through the Bonferroni correction at *p*<1.93×10^−5^ for the amino acids and *p*<4.35×10^−4^ for the classical HLA alleles.

## RESULTS

The study tested the association with CAP in a total of 7,638,472 variants from 3,508 controls and 257 patients (**Figure 1**), in which *S. pneumoniae* was identified in 30% of cases, and the remaining 70% were patients where the causative microorganism was not identified. The clinical and demographic characteristics of the study subjects are shown in **Supplementary Table 1**. Overall, we did not detect inflation of the association results as the lambda of the study barely deviated from the expected under the null (λ=1.04). Association testing revealed a total of 67 genome-wide significant variants (**Supplementary Table S2**) which were located on chromosomes 4q28.2, 6p21.32, 11p12, and 20q11.22. Regional association plots for these results are provided in the **Supplementary Figure S1**.

**Figure 1.**
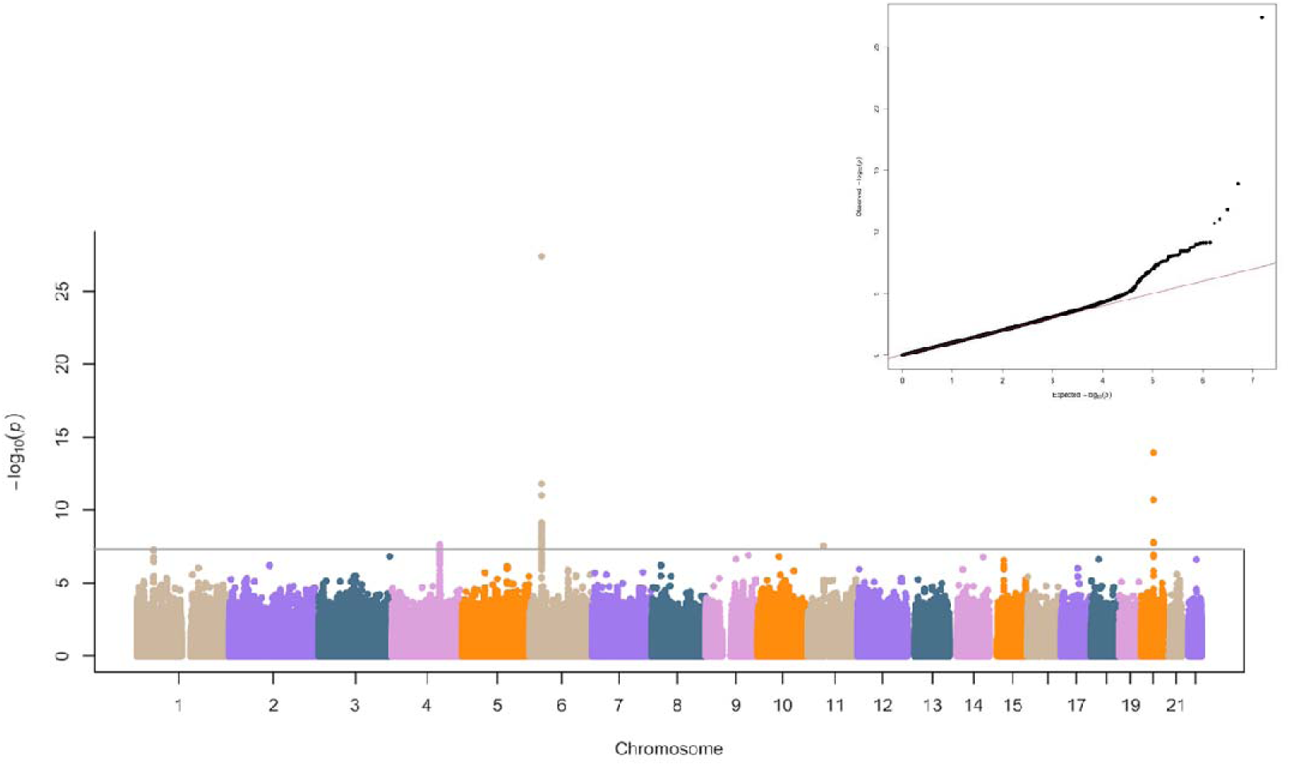
Manhattan plot of the genome-wide association study results for adult CAP. The x-axis represents the chromosome positions while the y-axis represents the transformed *p*-values (–log_10_ *p*-value). The horizontal line represents the genome-wide significance threshold (*p*-value=5.0×10^−8^). The inset of the top right represents the Quantile-Quantile (QQ) plot with the observed (y-axis) *vs.* the expected (x-axis) −log_10_ *p*-values of the association study (inflation λ= 1.04).

There were six independently associated sentinel variants (**Table 1**): rs34955650 at 4q28.2 (*p*=2.41×10^−8^) intergenic to *C4orf33* and LINC02466; three at 6p21.32, rs456261 and rs2076775 that are both intronic to *PFDN6* (*p*=4.00×10^−28^) and *SYNGAP1* (*p*=7.26×10^−10^), respectively, and rs213226 that is intergenic to *RING1* and *HCG25* (*p*=8.64×10^−9^); rs117203606 at 11p12 (*p*=2.90×10^−8^) which is intergenic to LINC02740 and *HNRNPKP3*; and rs45577437 at 20q11.22 (*p*=1.21×10^−14^) that is exonic to *ZNF341*. The latter locates in exon 5 of *ZNF341* and predicts a missense variant from a nonpolar to a polar uncharged amino acid at position 185 of the protein (p.Pro185Ser). The functional annotation suggests that rs45577437 may have a biological impact based on its high CADD score (CADD=17.92), despite it is lower than the MSC for this gene (MSC=31.62). Models adjusting for other covariates did not substantially modify these results (**Supplementary Table S3**).

**Table 1.** Results of the sentinel variants independently associated with CAP.

| Variant | Position | p-value | MAF | OR [95% CI] | A1/A2 | Func.Gene | Nearest gene(s) |
| --- | --- | --- | --- | --- | --- | --- | --- |
| rs34955650 | 4:130254264 | 2.41x10<sup>-8</sup> | 0.05 | 2.46 [1.80-3.38] | C/T | intergenic | *C4orf33*, *LINC02466* |
| rs213226 | 6:33209310 | 8.64x10<sup>-9</sup> | 0.17 | 1.84 [1.49-2.26] | A/G | intergenic | *RING1*, *HCG25* |
| rs456261 | 6:33258443 | 4.00x10<sup>-28</sup> | 0.02 | 8.23 [5.65-11.99] | G/A | intronic | *PFDN6* |
| rs2076775 | 6:33394253 | 7.26x10<sup>-10</sup> | 0.14 | 0.50 [0.41-0.63] | C/G | intronic | *SYNGAP1* |
| rs117203606 | 11:42330132 | 2.90x10<sup>-8</sup> | 0.11 | 0.21 [0.12-0.36] | G/A | intergenic | *LINC02740*,<br>*HNRNPKP3* |
| rs45577437 | 20:32341041 | 1.21x10<sup>-14</sup> | 0.44 | 0.46 [0.37-0.56] | C/T | exonic | *ZNF341* |

Position: chromosome and base pair according to GRCh37/hg19; MAF: minor allele frequency in the study population; OR: odds ratio; CI: confidence interval; A1: Non-effect allele; A2: Effect allele; Func.Gene: gene location.

The results for the association of the two severe pneumonia outcomes considered by separate or combined (i.e., severe sepsis or septic shock and severe respiratory insufficiency) for the six independently sentinel variants are shown in **Supplementary Table S4**. For these sub-analyses, despite the low sample size reduction, all variants reach significance at the nominal level and the direction of effect is still maintained.

Bayesian fine mapping around each of the four chromosome loci to identify the most likely causal variants driving the association was unable to prioritize variants for 11p12 and 20q11.22. However, it was able to delineate a credible set of 52 variants for 4q28.2 and 25 variants for 6p21.32 (**Supplementary Table S5** and **Figure S2**). The variant from each credible set with the highest V2G score was used to assign the most likely gene involved in the association. At 6p21.32, the ranking prioritized the *TAPBP* gene (V2G max. score=0.41 and MSC=4.26), encoding the TAP binding protein. At 4q28.2, the *C4orf33* gene was prioritized (V2G max. score=0.15 and MSC=4.87). For downstream functional analysis, we selected those genetic variants with the highest probability of biological effect based on the CADD score from the delineated credible sets in the associated loci. For the 6p21.32 locus, we assessed 10 variants (CADD>MSC), and the predictions suggest a relevant biological impact since some of these SNPs predict the affectation of transcription factor binding and regulatory motif and DNase I hypersensitive sites. Furthermore, rs381847, rs2247385, and rs456261 may affect enhancer (H3K4me1 and H3K27ac) and promoter (H3K4me3 and H3K9ac) histone marks in several cell lines, including lung tissue and immune cells. Moreover, these 10 variants are eQTLs of immunity genes, including HLA genes and *TAPBP* (**Supplementary Table S6**). For the credible set of 4q28.2, we selected 13 variants with CADD>MSC. Two of these are rs17014611 and rs35004602, both linked to predictions of enhancer histone marks affectation in lung and immune cell lines, and DNase I hypersensitive sites in fetal lung (**Supplementary Table S7**).

According to GTEx, the highest *TAPBP* gene expression was detected on spleen, lungs, lymphocytes, and whole blood. In fact, the three independent sentinel variants of 6p21.32 were eQTLs for the *TAPBP* gene on artery, esophagus, lung, and whole blood (**Supplementary Table S8**). No significant eQTLs were found for any tissue at GTEx for the other three independent sentinel variants at chromosomes 4q28.2, 11p12, and 20q11.22 (rs34955650, rs117203606, and rs45577437). Furthermore, we found a significant upregulation of *TAPBP* (lowest *p*=1.44×10^−6^ among the four probe sets available) and for *C4orf33* (*p*=9.67×10^−5^) among ICU patients with CAP compared to controls (GEO: GSE65682). *ZNF341* did not show a significant gene expression difference (**Supplementary Figure S3**).

Finally, we assessed the association of 155 classical HLA alleles and the 2,584 amino acids for eight classical HLA genes with CAP. Despite the key implication of the HLA genes in infectious diseases, all associations tested were found non-significant (**Supplementary Figure S4**, and **Table S9**).

## DISCUSSION

Here, we describe the results of a GWAS of CAP conducted in Spanish population. We identified six independently associated variants from four chromosome loci (4q28.2, 6p21.32, 11p12, and 20q11.22) reaching genome-wide significance. This GWAS focusing on only CAP patients, enriched for pneumococcus as causative etiology, has allowed to identify novel loci associated with CAP susceptibility. Mutations at two of these loci, *ZNF341* and *TAPBP*, cause previously known IEIs involved in the susceptibility to bacterial infections, particularly pneumonia.

One of the independent variants detected is located on 4q28.2, intergenic to *C4orf33* and a long non-coding RNA (LINC02466) which has been involved in cancer (Li et al., 2021). Besides, the sentinel variants on chromosome 6 have been previously associated with platelet and blood cell count, type-I diabetes, or celiac disease, among other traits (GWAS data available at Open Target Genetics). The most significant variant, rs456261, is intronic to the *PFDN6* gene, which encodes a member of the beta subunit of the prefoldin complex involved in protein folding and implicated in tumorigenesis (Liang et al., 2020). Gao and colleagues reported that this gene may be involved in susceptibility to COVID-19 in patients with lung adenocarcinoma and associated with mortality (Gao et al., 2021). Another intronic variant was detected at *SYNGAP1*, which encodes for the synaptic Ras GTPase activating protein 1 which has been associated with neurodevelopmental disorders (Llamosas et al., 2020). Finally, another variant was prioritized in 11p12 intergenic to a long non-coding RNA and a pseudogene (*HNRNPKP3*). To our knowledge, these genes have not been associated with prior predisposition or severity of infections. However, further studies of functional characterization are needed to assess the biological effects of the identified genes and variants in pneumonia.

The sentinel variant of the 20q11.22 locus observed in this study is located in exon 5 of *ZNF341* and predicts a missense change (p.Pro185Ser). ZNF341 acts as a DNA-binding transcription factor, primarily as an activator of Signal Transducer and Activator of Transcription *STAT3* gene, and, to a lesser extent, a number of other genes such as *STAT1* (Asano et al., 2021; August, 2018; Béziat et al., 2018; Frey-Jakobs et al., 2018). STAT3 plays a critical role in responses to many cytokines as well as to several growth factors. Autosomal dominant (AD) mutations at *STAT3* are the main cause of hyper-immunoglobulin E syndrome (HIES). HIES is characterized by elevated serum IgE levels, recurrent bacterial and candida infections, eczema with cold staphylococcal skin abscesses, and other non-immunologic features that affect the skeleton, dentition, and connective tissue. HIES patients have recurrent pneumonia caused by *S. aureus*, *S. pneumoniae*, or *Haemophilus influenzae*, and the pulmonary recovery can involve abnormalities characterized by bronchiectasis and pneumatoceles (Minegishi, 2021, 2023; Tsilifis et al., 2021). HIES caused by ZNF341 deficiency has been described as a phenocopy of HIES due to *STAT3* mutations, although with fewer extrahematopoietic manifestations (Béziat et al., 2018; Frey-Jakobs et al., 2018; Minegishi, 2021, 2023; Tsilifis et al., 2021). In addition, the lymphocyte subsets of ZNF341-deficient patients closely resemble that of STAT3-HIES, including decreased Th17 cells, T follicular helper cells, and memory B cell deficiency (Béziat et al., 2018; Frey-Jakobs et al., 2018; Minegishi, 2021, 2023). The susceptibility to pneumonia by pneumococcus and *H. influenzae* in patients with STAT3- and ZNF341-HIES is caused by the defective development of B cells and antibody production (Béziat et al., 2018; Frey-Jakobs et al., 2018; Tsilifis et al., 2021). Based on this evidence, variants affecting the function of *ZNF341* could also play an important role in CAP and its severity.

The study also prioritized possibly damaging variants in the *TAPBP* gene that were eQTLs for that gene in different tissues. It also revealed that *TAPBP* gene expression was upregulated among ICU patients with CAP compared to controls, although the role of profound inflammatory dysregulation in critical patients cannot be discarded. The *TAPBP* gene encodes the transporter associated with antigen processing (TAP) binding protein, also called tapasin. Tapasin is part of the peptide loading complex (PLC), which coordinates loading of high affinity peptides onto nascent HLA class I (HLA-I) molecules (Blees et al., 2017; Grandea et al., 2000; Grandea & Van Kaer, 2001; Jiang et al., 2022; Mantel et al., 2022; Peh et al., 1998; Pishesha et al., 2022). Tapasin has a central role in the classical pathway of HLA-I presentation of endogenous peptides and in cross-presentation of exogenous antigens by professional antigen presenting cells (Mantel et al., 2022). Multiple HLA-I molecules have been reported to be tapasin-dependent, among them the *HLA-B*08:01* allele (Rizvi et al., 2014). In spite that a previous GWAS found this class I HLA allele associated with pneumonia susceptibility (Tian et al., 2017), *HLA-B*08:01* was not associated with CAP in our study (*p*=0.45). Gene defects in *TAPBP* or in other PLC components cause an extremely rare IEI known as bare lymphocyte syndrome type I (BLS-I) (Darazam et al., 2023; Hanalioglu et al., 2017; Hanna & Etzioni, 2014; Shrestha et al., 2012; Su & Lenardo, 2014; Yabe et al., 2002). Despite the clinical and biological heterogeneity, BLS-I patients usually develop symptoms well into late childhood, although some patients remain asymptomatic even in adulthood. They are characterized by recurrent respiratory tract infections, mostly by *S. pneumoniae* and *H. influenzae*, associated with the development of bronchiectasis and respiratory insufficiency, as well as to cutaneous manifestations (Darazam et al., 2023; Hanna & Etzioni, 2014; Shrestha et al., 2012; Yabe et al., 2002; Zimmer et al., 2005). It is not clear how defects of HLA-I antigen presentation predispose to infections by extracellular bacteria. However, impaired antibody production (Kobrynski et al., 2005; Su & Lenardo, 2014; Yucesoy et al., 2013), a macrophage survival defect, and/or a defect of the elimination of bacteria by macrophages (Lapenna et al., 2017) may be involved.

We acknowledge some limitations of the study. First and foremost, the study was based on a small sample of cases. The increase in sample size will allow revealing more susceptibility loci beyond those identified here. Secondly, half of the patients had a low Pneumonia Severity Index. However, when the analyses only included the patients with severe sepsis or septic shock, or those with severe respiratory insufficiency, association results of the sentinel variants were maintained. Thirdly, our study lacks a formal assessment of the full spectrum of genetic variants (including other types of variation beyond SNPs and small INDELs, and rare variants) for which complementary approaches based on Next-Generation Sequencing, such as whole exome or genome sequencing, would be necessary. Fourth, the etiology of CAP was undetermined in 70% of the patients studied. Previous studies suggest that pneumococci cause most CAP cases in which negative test results were found using conventional microbiological methods, particularly in Spain (Gadsby et al., 2016; Johansson et al., 2010; Torres et al., 2018). However, infection by other bacteria and viruses is still expected. Some other common bacteria in CAP, such as *H. influenzae* or *Moraxella catharralis,* are also probably increased in the patients without confirmed microbial etiology (Gadsby et al., 2016; Johansson et al., 2010; Musher et al., 2017; Shoar et al., 2021). These bacteria are also frequently observed in IEI predisposing to pneumococcal pneumonia, such as in HIES and TAPBP deficiency (Hanna & Etzioni, 2014; Shrestha et al., 2012; Zimmer et al., 2005). In addition, a substantial proportion of patients with viral infections present with bacterial coinfection, particularly pneumococcus (Gadsby et al., 2016; Hedberg et al., 2022; Johansson et al., 2010; Liu et al., 2023; Musher et al., 2017). Finally, we used controls that could have introduced some bias in the results since, despite hospitalization for CAP was not recorded for these donors during the recruitment, we cannot discard that they could develop it during their lifetime. These types of controls, however, are widely used in genetic studies for multiple infectious diseases by convenience and by providing equivalent results as if controls comprise mild or asymptomatic patients (Chen et al., 2021; Cruz et al., 2022; Degenhardt et al., 2022).

## CONCLUSION

In summary, we report four novel loci associated with CAP, including two genes that were previously known to cause IEIs predisposing to bacterial pneumonia. Complementary studies are required to better define the mechanistic links of these variants and genes on predisposition to pneumonia.

## Data Availability

All data produced in the present study are available upon reasonable request to the authors

## REFERENCES

Asano, T., Khourieh, J., Zhang, P., Rapaport, F., Spaan, A. N., Li, J., Lei, W.-T., Pelham, S. J., Hum, D., Chrabieh, M., Han, J. E., Guérin, A., Mackie, J., Gupta, S., Saikia, B., Baghdadi, J. E. I., Fadil, I., Bousfiha, A., Habib, T., … Boisson, B. (2021). Human STAT3 variants underlie autosomal dominant hyper-IgE syndrome by negative dominance. Journal of Experimental Medicine, 218(8). 10.1084/jem.20202592

August, A. (2018). Who regulates whom: ZNF341 is an additional player in the STAT3/T H 17 song. Science Immunology, 3(24). doi: 10.1126/sciimmunol.aat9779

Béziat, V., Li, J., Lin, J.-X., Ma, C. S., Li, P., Bousfiha, A., Pellier, I., Zoghi, S., Baris, S., Keles, S., Gray, P., Du, N., Wang, Y., Zerbib, Y., Lévy, R., Leclercq, T., About, F., Lim, A. I., Rao, G., … Casanova, J.-L. (2018). A recessive form of hyper-IgE syndrome by disruption of ZNF341-dependent STAT3 transcription and activity. Science Immunology, 3(24). 10.1126/sciimmunol.aat4956

Bjarnason, A., Westin, J., Lindh, M., Andersson, L.-M., Kristinsson, K. G., Löve, A., Baldursson, O., & Gottfredsson, M. (2018). Incidence, Etiology, and Outcomes of Community-Acquired Pneumonia: A Population-Based Study. Open Forum Infectious Diseases, 5(2). 10.1093/ofid/ofy010

Blees, A., Januliene, D., Hofmann, T., Koller, N., Schmidt, C., Trowitzsch, S., Moeller, A., & Tampé, R. (2017). Structure of the human MHC-I peptide-loading complex. Nature, 551(7681), 525–528. 10.1038/nature24627

Boisson, B. (2020). The genetic basis of pneumococcal and staphylococcal infections: inborn errors of human TLR and IL-1R immunity. Human Genetics, 139(6–7), 981–991. 10.1007/s00439-020-02111-z

Bone, R. C., Balk, R. A., Cerra, F. B., Dellinger, R. P., Fein, A. M., Knaus, W. A., Schein, R. M. H., & Sibbald, W. J. (1992). Definitions for Sepsis and Organ Failure and Guidelines for the Use of Innovative Therapies in Sepsis. Chest, 101(6), 1644–1655. 10.1378/chest.101.6.1644

Boyle, A. P., Hong, E. L., Hariharan, M., Cheng, Y., Schaub, M. A., Kasowski, M., Karczewski, K. J., Park, J., Hitz, B. C., Weng, S., Cherry, J. M., & Snyder, M. (2012). Annotation of functional variation in personal genomes using RegulomeDB. Genome Research, 22(9), 1790–1797. 10.1101/gr.137323.112

Casanova, J.-L., & Abel, L. (2021). Lethal Infectious Diseases as Inborn Errors of Immunity: Toward a Synthesis of the Germ and Genetic Theories. Annual Review of Pathology: Mechanisms of Disease, 16(1), 23–50. 10.1146/annurev-pathol-031920-101429

Casanova, J.-L., & Abel, L. (2022). From rare disorders of immunity to common determinants of infection: Following the mechanistic thread. Cell, 185(17), 3086–3103. 10.1016/j.cell.2022.07.004

Chang, C. C., Chow, C. C., Tellier, L. C., Vattikuti, S., Purcell, S. M., & Lee, J. J. (2015). Second-generation PLINK: rising to the challenge of larger and richer datasets. GigaScience, 4(1), 7. 10.1186/s13742-015-0047-8

Chen, H.-H., Shaw, D. M., Petty, L. E., Graff, M., Bohlender, R. J., Polikowsky, H. G., Zhong, X., Kim, D., Buchanan, V. L., Preuss, M. H., Shuey, M. M., Loos, R. J. F., Huff, C. D., Cox, N. J., Bastarache, J. A., Bastarache, L., North, K. E., & Below, J. E. (2021). Host genetic effects in pneumonia. The American Journal of Human Genetics, 108(1), 194–201. 10.1016/j.ajhg.2020.12.010

Cruz, R., Diz-de Almeida, S., López de Heredia, M., Quintela, I., Ceballos, F. C., Pita, G., Lorenzo-Salazar, J. M., González-Montelongo, R., Gago-Domínguez, M., Sevilla Porras, M., Tenorio Castaño, J. A., Nevado, J., Aguado, J. M., Aguilar, C., Aguilera-Albesa, S., Almadana, V., Almoguera, B., Alvarez, N., Andreu-Bernabeu, Á., … Carracedo, A. (2022). Novel genes and sex differences in COVID-19 severity. Human Molecular Genetics, 31(22), 3789–3806. 10.1093/hmg/ddac132

Darazam, I. A., Hakamifard, A., Momenilandi, M., Materna, M., Gharehbagh, F. J., Shahrooei, M., Olyaei, N. A., Zerehpoosh, F. B., Fayand, A., Hatami, F., Lotfollahi, L., Mansouri, N., Casanova, J.-L., Béziat, V., & Mansouri, D. (2023). Delayed Diagnosis of Chronic Necrotizing Granulomatous Skin Lesions due to TAP2 Deficiency. Journal of Clinical Immunology, 43(1), 217–228. 10.1007/s10875-022-01374-7

Degenhardt, F., Ellinghaus, D., Juzenas, S., Lerga-Jaso, J., Wendorff, M., Maya-Miles, D., Uellendahl-Werth, F., ElAbd, H., Rühlemann, M. C., Arora, J., Özer, O., Lenning, O. B., Myhre, R., Vadla, M. S., Wacker, E. M., Wienbrandt, L., Blandino Ortiz, A., de Salazar, A., Garrido Chercoles, A., … Franke, A. (2022). Detailed stratified GWAS analysis for severe COVID-19 in four European populations. Human Molecular Genetics, 31(23), 3945–3966. 10.1093/hmg/ddac158

Dion, C. F., & Ashurst, J. V. (2022). Streptococcus Pneumoniae. Available from: https://www.ncbi.nlm.nih.gov/books/NBK470537/

Frey-Jakobs, S., Hartberger, J. M., Fliegauf, M., Bossen, C., Wehmeyer, M. L., Neubauer, J. C., Bulashevska, A., Proietti, M., Fröbel, P., Nöltner, C., Yang, L., Rojas-Restrepo, J., Langer, N., Winzer, S., Engelhardt, K. R., Glocker, C., Pfeifer, D., Klein, A., Schäffer, A. A., … Grimbacher, B. (2018). ZNF341 controls STAT3 expression and thereby immunocompetence. Science Immunology, 3(24). 10.1126/sciimmunol.aat4941

Gadsby, N. J., Russell, C. D., McHugh, M. P., Mark, H., Conway Morris, A., Laurenson, I. F., Hill, A. T., & Templeton, K. E. (2016). Comprehensive Molecular Testing for Respiratory Pathogens in Community-Acquired Pneumonia. Clinical Infectious Diseases, 62(7), 817–823. 10.1093/cid/civ1214

Gallagher, P. M., Lowe, G., Fitzgerald, T., Bella, A., Greene, C. M., McElvaney, N. G., & O’Neill, S. J. (2003). Association of IL-10 polymorphism with severity of illness in community acquired pneumonia. Thorax, 58(2), 154–156. 10.1136/thorax.58.2.154

Gao, L., Li, G.-S., Li, J.-D., He, J., Zhang, Y., Zhou, H.-F., Kong, J.-L., & Chen, G. (2021). Identification of the susceptibility genes for COVID-19 in lung adenocarcinoma with global data and biological computation methods. Computational and Structural Biotechnology Journal, 19, 6229–6239. 10.1016/j.csbj.2021.11.026

García-Laorden, M. I., Hernández-Brito, E., Muñoz-Almagro, C., Pavlovic-Nesic, S., Rúa-Figueroa, I., Briones, M. L., Rajas, O., Borderías, L., Payeras, A., Lorente, L., Freixinet, J., Ferreres, J., Obando, I., González-Quevedo, N., Rodríguez de Castro, F., Solé-Violán, J., & Rodríguez-Gallego, C. (2020). Should MASP-2 Deficiency Be Considered a Primary Immunodeficiency? Relevance of the Lectin Pathway. Journal of Clinical Immunology, 40(1), 203–210. 10.1007/s10875-019-00714-4

García-Laorden, M. I., Rodríguez de Castro, F., Solé-Violán, J., Rajas, O., Blanquer, J., Borderías, L., Aspa, J., Briones, M. L., Saavedra, P., Marcos-Ramos, J. A., González-Quevedo, N., Sologuren, I., Herrera-Ramos, E., Ferrer, J. M., Rello, J., & Rodríguez-Gallego, C. (2011). Influence of genetic variability at the surfactant proteins A and D in community-acquired pneumonia: a prospective, observational, genetic study. Critical Care, 15(1), R57. 10.1186/cc10030

Garcia-Laorden, M. I., Sole-Violan, J., de Castro, F. R., Aspa, J., Briones, M. L., Garcia-Saavedra, A., Rajas, O., Blanquer, J., Caballero-Hidalgo, A., Marcos-Ramos, J. A., Hernandez-Lopez, J., & Rodriguez-Gallego, C. (2008). Mannose-binding lectin and mannose-binding lectin–associated serine protease 2 in susceptibility, severity, and outcome of pneumonia in adults. Journal of Allergy and Clinical Immunology, 122(2), 368–374.e2. 10.1016/j.jaci.2008.05.037

Ghoussaini, M., Mountjoy, E., Carmona, M., Peat, G., Schmidt, E. M., Hercules, A., Fumis, L., Miranda, A., Carvalho-Silva, D., Buniello, A., Burdett, T., Hayhurst, J., Baker, J., Ferrer, J., Gonzalez-Uriarte, A., Jupp, S., Karim, M. A., Koscielny, G., Machlitt-Northen, S., … Dunham, I. (2021). Open Targets Genetics: systematic identification of trait-associated genes using large-scale genetics and functional genomics. Nucleic Acids Research, 49(D1), D1311–D1320. 10.1093/nar/gkaa840

Grandea, A. G., Golovina, T. N., Hamilton, S. E., Sriram, V., Spies, T., Brutkiewicz, R. R., Harty, J. T., Eisenlohr, L. C., & Van Kaer, L. (2000). Impaired Assembly yet Normal Trafficking of MHC Class I Molecules in Tapasin Mutant Mice. Immunity, 13(2), 213–222. 10.1016/S1074-7613(00)00021-2

Grandea, A. G., & Van Kaer, L. (2001). Tapasin: an ER chaperone that controls MHC class I assembly with peptide. Trends in Immunology, 22(4), 194–199. 10.1016/S1471-4906(01)01861-0

Hanalioglu, D., Ayvaz, D. C., Ozgur, T. T., van der Burg, M., Sanal, O., & Tezcan, I. (2017). A novel mutation in TAP1 gene leading to MHC class I deficiency: Report of two cases and review of the literature. Clinical Immunology, 178, 74–78. 10.1016/j.clim.2017.01.011

Hanna, S., & Etzioni, A. (2014). MHC class I and II deficiencies. Journal of Allergy and Clinical Immunology, 134(2), 269–275. 10.1016/j.jaci.2014.06.001

Hedberg, P., Johansson, N., Ternhag, A., Abdel-Halim, L., Hedlund, J., & Nauclér, P. (2022). Bacterial co-infections in community-acquired pneumonia caused by SARS-CoV-2, influenza virus and respiratory syncytial virus. BMC Infectious Diseases, 22(1), 108. 10.1186/s12879-022-07089-9

Hutchinson, A., Watson, H., & Wallace, C. (2020). Improving the coverage of credible sets in Bayesian genetic fine-mapping. PLOS Computational Biology, 16(4), e1007829. 10.1371/journal.pcbi.1007829

Ishak, A., Mehendale, M., AlRawashdeh, M. M., Sestacovschi, C., Sharath, M., Pandav, K., & Marzban, S. (2022). The association of COVID-19 severity and susceptibility and genetic risk factors: A systematic review of the literature. Gene, 836, 146674. 10.1016/j.gene.2022.146674

Itan, Y., Shang, L., Boisson, B., Ciancanelli, M. J., Markle, J. G., Martinez-Barricarte, R., Scott, E., Shah, I., Stenson, P. D., Gleeson, J., Cooper, D. N., Quintana-Murci, L., Zhang, S.-Y., Abel, L., & Casanova, J.-L. (2016). The mutation significance cutoff: gene-level thresholds for variant predictions. Nature Methods, 13(2), 109–110. 10.1038/nmeth.3739

Jiang, J., Taylor, D. K., Kim, E. J., Boyd, L. F., Ahmad, J., Mage, M. G., Truong, H. V., Woodward, C. H., Sgourakis, N. G., Cresswell, P., Margulies, D. H., & Natarajan, K. (2022). Structural mechanism of tapasin-mediated MHC-I peptide loading in antigen presentation. Nature Communications, 13(1), 5470. 10.1038/s41467-022-33153-8

Johansson, N., Kalin, M., Tiveljung-Lindell, A., Giske, C. G., & Hedlund, J. (2010). Etiology of Community-Acquired Pneumonia: Increased Microbiological Yield with New Diagnostic Methods. Clinical Infectious Diseases, 50(2), 202–209. 10.1086/648678

Kang HM, Canouil M, Nguyen P. (2022). EPACTS (Efficient and Parallelizable Association container toolbox)

Kloek, A. T., Brouwer, M. C., & van de Beek, D. (2019). Host genetic variability and pneumococcal disease: a systematic review and meta-analysis. BMC Medical Genomics, 12(1), 130. 10.1186/s12920-019-0572-x

Kobrynski, L. J., Sousa, A. O., Nahmias, A. J., & Lee, F. K. (2005). Cutting Edge: Antibody Production to Pneumococcal Polysaccharides Requires CD1 Molecules and CD8+ T Cells. The Journal of Immunology, 174(4), 1787–1790. 10.4049/jimmunol.174.4.1787

Lanks, C. W., Musani, A. I., & Hsia, D. W. (2019). Community-acquired Pneumonia and Hospital-acquired Pneumonia. Medical Clinics of North America, 103(3), 487–501. 10.1016/j.mcna.2018.12.008

Lapenna, A., Omar, I., & Berger, M. (2017). A novel spontaneous mutation in the TAP2 gene unravels its role in macrophage survival. Immunology, 150(4), 432–443. 10.1111/imm.12694

Lees, J. A., Ferwerda, B., Kremer, P. H. C., Wheeler, N. E., Serón, M. V., Croucher, N. J., Gladstone, R. A., Bootsma, H. J., Rots, N. Y., Wijmega-Monsuur, A. J., Sanders, E. A. M., Trzciński, K., Wyllie, A. L., Zwinderman, A. H., van den Berg, L. H., van Rheenen, W., Veldink, J. H., Harboe, Z. B., Lundbo, L. F., … van de Beek, D. (2019). Joint sequencing of human and pathogen genomes reveals the genetics of pneumococcal meningitis. Nature Communications, 10(1), 2176. 10.1038/s41467-019-09976-3

Li, Y., Wei, W., An, S., Jiang, J., He, J., Zhang, H., Wang, G., Han, J., Liang, B., Ye, L., & Liang, H. (2021). Identification and analysis of lncRNA, microRNA and mRNA expression profiles and construction of ceRNA network in Talaromyces marneffei-infected THP-1 macrophage. PeerJ, 9, e10529. 10.7717/peerj.10529

Liang, J., Xia, L., Oyang, L., Lin, J., Tan, S., Yi, P., Han, Y., Luo, X., Wang, H., Tang, L., Pan, Q., Tian, Y., Rao, S., Su, M., Shi, Y., Cao, D., Zhou, Y., & Liao, Q. (2020). The functions and mechanisms of prefoldin complex and prefoldin-subunits. Cell & Bioscience, 10(1), 87. 10.1186/s13578-020-00446-8

Liu, Y.-N., Zhang, Y.-F., Xu, Q., Qiu, Y., Lu, Q.-B., Wang, T., Zhang, X.-A., Lin, S.-H., Lv, C.-L., Jiang, B.-G., Li, H., Li, Z.-J., Gao, G. F., Yang, W.-Z., Hay, S. I., Wang, L.-P., Fang, L.-Q., & Liu, W. (2023). Infection and co-infection patterns of community-acquired pneumonia in patients of different ages in China from 2009 to 2020: a national surveillance study. The Lancet Microbe, 4(5), e330–e339. 10.1016/S2666-5247(23)00031-9

Llamosas, N., Arora, V., Vij, R., Kilinc, M., Bijoch, L., Rojas, C., Reich, A., Sridharan, B., Willems, E., Piper, D. R., Scampavia, L., Spicer, T. P., Miller, C. A., Holder, J. L., & Rumbaugh, G. (2020). SYNGAP1 Controls the Maturation of Dendrites, Synaptic Function, and Network Activity in Developing Human Neurons. The Journal of Neuroscience, 40(41), 7980–7994. 10.1523/JNEUROSCI.1367-20.2020

Loh, P.-R., Danecek, P., Palamara, P. F., Fuchsberger, C., A Reshef, Y., K Finucane, H., Schoenherr, S., Forer, L., McCarthy, S., Abecasis, G. R., Durbin, R., & L Price, A. (2016). Reference-based phasing using the Haplotype Reference Consortium panel. Nature Genetics, 48(11), 1443–1448. 10.1038/ng.3679

Luo, Y., Kanai, M., Choi, W., Li, X., Sakaue, S., Yamamoto, K., Ogawa, K., Gutierrez-Arcelus, M., Gregersen, P. K., Stuart, P. E., Elder, J. T., Forer, L., Schönherr, S., Fuchsberger, C., Smith, A. V., Fellay, J., Carrington, M., Haas, D. W., Guo, X., … Raychaudhuri, S. (2021). A high-resolution HLA reference panel capturing global population diversity enables multi-ancestry fine-mapping in HIV host response. Nature Genetics, 53(10), 1504–1516. 10.1038/s41588-021-00935-7

Mantel, I., Sadiq, B. A., & Blander, J. M. (2022). Spotlight on TAP and its vital role in antigen presentation and cross-presentation. Molecular Immunology, 142, 105–119. 10.1016/j.molimm.2021.12.013

McCarthy, S., Das, S., Kretzschmar, W., Delaneau, O., Wood, A. R., Teumer, A., Kang, H. M., Fuchsberger, C., Danecek, P., Sharp, K., Luo, Y., Sidore, C., Kwong, A., Timpson, N., Koskinen, S., Vrieze, S., Scott, L. J., Zhang, H., Mahajan, A., … Haplotype Reference Consortium. (2016). A reference panel of 64,976 haplotypes for genotype imputation. Nature Genetics, 48(10), 1279–1283. 10.1038/ng.3643

Millett, E. R. C., Quint, J. K., Smeeth, L., Daniel, R. M., & Thomas, S. L. (2013). Incidence of Community-Acquired Lower Respiratory Tract Infections and Pneumonia among Older Adults in the United Kingdom: A Population-Based Study. PLoS ONE, 8(9), e75131. 10.1371/journal.pone.0075131

Minegishi, Y. (2021). Hyper-IgE syndrome, 2021 update. Allergology International, 70(4), 407–414. 10.1016/j.alit.2021.07.007

Minegishi, Y. (2023). The signal transducer and activator of transcription 3 at the center of the causative gene network of the hyper-IgE syndrome. Current Opinion in Immunology, 80, 102264. 10.1016/j.coi.2022.102264

Mortensen, E., & Metersky, M. (2012). Long-Term Mortality after Pneumonia. Seminars in Respiratory and Critical Care Medicine, 33(03), 319–324. 10.1055/s-0032-1315644

Musher, D. M., Abers, M. S., & Bartlett, J. G. (2017). Evolving Understanding of the Causes of Pneumonia in Adults, With Special Attention to the Role of Pneumococcus. Clinical Infectious Diseases, 65(10), 1736–1744. 10.1093/cid/cix549

Peh, C. A., Burrows, S. R., Barnden, M., Khanna, R., Cresswell, P., Moss, D. J., & McCluskey, J. (1998). HLA-B27–Restricted Antigen Presentation in the Absence of Tapasin Reveals Polymorphism in Mechanisms of HLA Class I Peptide Loading. Immunity, 8(5), 531–542. 10.1016/S1074-7613(00)80558-0

Picard, C., Puel, A., Bustamante, J., Ku, C.-L., & Casanova, J.-L. (2003). Primary immunodeficiencies associated with pneumococcal disease. Current Opinion in Allergy and Clinical Immunology, 3(6), 451–459. 10.1097/00130832-200312000-00006

Pick, H., Daniel, P., Rodrigo, C., Bewick, T., Ashton, D., Lawrence, H., Baskaran, V., Edwards-Pritchard, R. C., Sheppard, C., Eletu, S. D., Rose, S., Litt, D., Fry, N. K., Ladhani, S., Chand, M., Trotter, C., McKeever, T. M., & Lim, W. S. (2020). Pneumococcal serotype trends, surveillance and risk factors in UK adult pneumonia, 2013–18. Thorax, 75(1), 38–49. 10.1136/thoraxjnl-2019-213725

Pishesha, N., Harmand, T. J., & Ploegh, H. L. (2022). A guide to antigen processing and presentation. Nature Reviews Immunology, 22(12), 751–764. 10.1038/s41577-022-00707-2

R Core Team. (2022). R: A Language and Environment for Statistical Computing. R Foundation for Statistical Computing.

Ramirez, J. A., Wiemken, T. L., Peyrani, P., Arnold, F. W., Kelley, R., Mattingly, W. A., Nakamatsu, R., Pena, S., Guinn, B. E., Furmanek, S. P., Persaud, A. K., Raghuram, A., Fernandez, F., Beavin, L., Bosson, R., Fernandez-Botran, R., Cavallazzi, R., Bordon, J., Valdivieso, C., … Carrico, R. M. (2017). Adults Hospitalized With Pneumonia in the United States: Incidence, Epidemiology, and Mortality. Clinical Infectious Diseases, 65(11), 1806–1812. 10.1093/cid/cix647

Ranzani, O. T., Prina, E., Menéndez, R., Ceccato, A., Cilloniz, C., Méndez, R., Gabarrus, A., Barbeta, E., Bassi, G. L., Ferrer, M., & Torres, A. (2017). New Sepsis Definition (Sepsis-3) and Community-acquired Pneumonia Mortality. A Validation and Clinical Decision-Making Study. American Journal of Respiratory and Critical Care Medicine, 196(10), 1287–1297. 10.1164/rccm.201611-2262OC

Reay, W. R., Geaghan, M. P., Agee, M., Alipanahi, B., Bell, R. K., Bryc, K., Elson, S. L., Fontanillas, P., Furlotte, N. A., Hicks, B., Hinds, D. A., Huber, K. E., Jewett, E. M., Jiang, Y., Kleinman, A., Lin, K.-H., Litterman, N. K., McCreight, J. C., McIntyre, M. H., … Cairns, M. J. (2022). The genetic architecture of pneumonia susceptibility implicates mucin biology and a relationship with psychiatric illness. Nature Communications, 13(1), 3756. 10.1038/s41467-022-31473-3

Rello, J., & Perez, A. (2016). Precision medicine for the treatment of severe pneumonia in intensive care. Expert Review of Respiratory Medicine, 10(3), 297–316. 10.1586/17476348.2016.1144477

Rivero-Calle, I., Pardo-Seco, J., Aldaz, P., Vargas, D. A., Mascarós, E., Redondo, E., Díaz-Maroto, J. L., Linares-Rufo, M., Fierro-Alacio, M. J., Gil, A., Molina, J., Ocaña, D., & Martinón-Torres, F. (2016). Incidence and risk factor prevalence of community-acquired pneumonia in adults in primary care in Spain (NEUMO-ES-RISK project). BMC Infectious Diseases, 16(1), 645. 10.1186/s12879-016-1974-4

Rizvi, S. M., Salam, N., Geng, J., Qi, Y., Bream, J. H., Duggal, P., Hussain, S. K., Martinson, J., Wolinsky, S. M., Carrington, M., & Raghavan, M. (2014). Distinct Assembly Profiles of HLA-B Molecules. The Journal of Immunology, 192(11), 4967–4976. 10.4049/jimmunol.1301670

Schaaf, B., Rupp, J., Müller-Steinhardt, M., Kruse, J., Boehmke, F., Maass, M., Zabel, P., & Dalhoff, K. (2005). The interleukin-6 −174 promoter polymorphism is associated with extrapulmonary bacterial dissemination in Streptococcus pneumoniae infection. Cytokine, 31(4), 324–328. 10.1016/j.cyto.2005.05.008

Scicluna, B. P., Klein Klouwenberg, P. M. C., van Vught, L. A., Wiewel, M. A., Ong, D. S. Y., Zwinderman, A. H., Franitza, M., Toliat, M. R., Nürnberg, P., Hoogendijk, A. J., Horn, J., Cremer, O. L., Schultz, M. J., Bonten, M. J., & van der Poll, T. (2015). A Molecular Biomarker to Diagnose Community-acquired Pneumonia on Intensive Care Unit Admission. American Journal of Respiratory and Critical Care Medicine, 192(7), 826–835. 10.1164/rccm.201502-0355OC

Shoar, S., Centeno, F. H., & Musher, D. M. (2021). Clinical Features and Outcomes of Community-Acquired Pneumonia Caused by Haemophilus influenzae. Open Forum Infectious Diseases, 8(4). 10.1093/ofid/ofaa622

Shrestha, D., Szöllősi, J., & Jenei, A. (2012). Bare lymphocyte syndrome: An opportunity to discover our immune system. Immunology Letters, 141(2), 147–157. 10.1016/j.imlet.2011.10.007

Solé-Violán, J., García-Laorden, M. I., Marcos-Ramos, J. A., Rodríguez de Castro, F., Rajas, O., Borderías, L., Briones, M. L., Herrera-Ramos, E., Blanquer, J., Aspa, J., Florido, Y., García-Bello, M. A., Ferrer-Agüero, J. M., Sologuren, I., & Rodriguez-Gallego, C. (2011). The Fcγ receptor IIA-H/H131 genotype is associated with bacteremia in pneumococcal community-acquired pneumonia*. Critical Care Medicine, 39(6), 1388–1393. 10.1097/CCM.0b013e31820eda74

Solé-Violán, J., Hernández-Brito, E., Valencia-Gallardo, J. M., Herrera-Ramos, E., Borderías, L., Briones, M. L., Rajas, O., Freixinet, J., Lorente, L., Payeras, A., Ferreres, J., Carbonell, N., González-Quevedo, N., González-Martín, J. M., Rodríguez de Castro, F., & Rodríguez-Gallego, C. (2021). Challenges in understanding host genetics and severity of community-acquired pneumonia. ERJ Open Research, 7(1), 00745–02020. 10.1183/23120541.00745-2020

Su, H. C., Jing, H., Zhang, Y., & Casanova, J.-L. (2023). Interfering with Interferons: A Critical Mechanism for Critical COVID-19 Pneumonia. Annual Review of Immunology, 41(1), 561–585. 10.1146/annurev-immunol-101921-050835

Su, H. C., & Lenardo, M. J. (2014). Combined Immune Deficiencies. In Stiehm’s Immune Deficiencies (pp. 143–169). Elsevier. 10.1016/B978-0-12-405546-9.00005-4

Theilacker, C., Sprenger, R., Leverkus, F., Walker, J., Häckl, D., von Eiff, C., & Schiffner-Rohe, J. (2021). Population-based incidence and mortality of community-acquired pneumonia in Germany. PLOS ONE, 16(6), e0253118. 10.1371/journal.pone.0253118

Tian, C., Hromatka, B. S., Kiefer, A. K., Eriksson, N., Noble, S. M., Tung, J. Y., & Hinds, D. A. (2017). Genome-wide association and HLA region fine-mapping studies identify susceptibility loci for multiple common infections. Nature Communications, 8(1), 599. 10.1038/s41467-017-00257-5

Torres, A., Cillóniz, C., Blasi, F., Chalmers, J. D., Gaillat, J., Dartois, N., Schmitt, H.-J., & Welte, T. (2018). Burden of pneumococcal community-acquired pneumonia in adults across Europe: A literature review. Respiratory Medicine, 137, 6–13. 10.1016/j.rmed.2018.02.007

Torres, A., Menéndez, R., España, P. P., Fernández-Villar, J. A., Marimón, J. M., Cilloniz, C., Méndez, R., Egurrola, M., Botana-Rial, M., Ercibengoa, M., Méndez, C., Cifuentes, I., Gessner, B. D., Torres, A., Cilloniz, C., Ceccato, A., San José, A., Bueno, L., Marco, F., … Samaniego, M. L. (2021). The Evolution and Distribution of Pneumococcal Serotypes in Adults Hospitalized With Community-Acquired Pneumonia in Spain Using a Serotype-Specific Urinary Antigen Detection Test: The CAPA Study, 2011–2018. Clinical Infectious Diseases, 73(6), 1075–1085. 10.1093/cid/ciab307

Tsilifis, C., Freeman, A. F., & Gennery, A. R. (2021). STAT3 Hyper-IgE Syndrome—an Update and Unanswered Questions. Journal of Clinical Immunology, 41(5), 864–880. 10.1007/s10875-021-01051-1

Ward, L. D., & Kellis, M. (2012). HaploReg: a resource for exploring chromatin states, conservation, and regulatory motif alterations within sets of genetically linked variants. Nucleic Acids Research, 40(D1), D930–D934. 10.1093/nar/gkr917

Welte, T., Torres, A., & Nathwani, D. (2012). Clinical and economic burden of community-acquired pneumonia among adults in Europe. Thorax, 67(1), 71–79. 10.1136/thx.2009.129502

Yabe, T., Kawamura, S., Sato, M., Kashiwase, K., Tanaka, H., Ishikawa, Y., Asao, Y., Oyama, J., Tsuruta, K., Tokunaga, K., Tadokoro, K., & Juji, T. (2002). A subject with a novel type I bare lymphocyte syndrome has tapasin deficiency due to deletion of 4 exons by Alu-mediated recombination. Blood, 100(4), 1496–1498. 10.1182/blood-2001-12-0252

Yucesoy, B., Talzhanov, Y., Johnson, V. J., Wilson, N. W., Biagini, R. E., Wang, W., Frye, B., Weissman, D. N., Germolec, D. R., Luster, M. I., & Barmada, M. M. (2013). Genetic variants within the MHC region are associated with immune responsiveness to childhood vaccinations. Vaccine, 31(46), 5381–5391. 10.1016/j.vaccine.2013.09.026

Zimmer, J., Andrès, E., Donato, L., Hanau, D., Hentges, F., & de la Salle, H. (2005). Clinical and immunological aspects of HLA class I deficiency. QJM: An International Journal of Medicine, 98(10), 719–727. 10.1093/qjmed/hci112

