## Supplementary material for "A genome-wide association study of adults with community-acquired pneumonia"

**Supplementary methods**

**Study design and cohort**

Patient inclusion criteria and phenotype descriptions

The patients with community-acquired pneumonia (CAP) included in this study were Spanish adult patients (older than 18 years) recruited between 2001 and 2016 from the following Spanish hospitals: Hospital Universitario de Gran Canaria Dr. Negrín (Gran Canaria), Hospital Universitario de Canarias (Tenerife), Hospital Clínico Universitario de Valencia (Valencia), Hospital Universitario de la Princesa (Madrid), Hospital General San Jorge (Zaragoza), and Hospital Universitario Son Llàtzer (Palma de Mallorca). Patients had not been hospitalized for at least the previous ten days and presented with symptoms of lower airway infection, together with the existence of a new radiological infiltrate and no alternative diagnoses during follow-up.

The etiology of pneumonia was presumptive if valid sputum samples (purulent sputum with more than 25 leucocytes and less than 10 epithelial cells per low magnification field) predominantly isolated *Streptococcus pneumoniae* with compatible Gram stain. The diagnosis was considered definitive in different cases: a) If there was a positive blood culture for *S. pneumonia*e; b) If *S. pneumoniae* was isolated from transthoracic puncture samples or pleural fluid; c) If there was positive urinary antigen for *S. pneumoniae* (using the Binax Now *S. pneumoniae* urinary Antigen Test); d) and if *S. pneumoniae* counts > 103 cfu/mL were found in telescoped catheter (TC) or lung biopsy (PB) samples, > 104 cfu/mL in bronchoalveolar lavage (BAL), or > 105 cfu/mL in tracheal aspirate samples.

**Gene expression dataset**

We accessed the gene expression dataset with accession GSE65682 from the Gene Expression Omnibus (GEO) public repository at the National Center for Biotechnology Information. The dataset corresponds to the study "Genome-wide blood transcriptional profiling in critically ill patients - MARS consortium" (Scicluna et al., 2015). Based on the provided information, we selected the data from patients who had been admitted to the ICU with a CAP diagnosis. Patients were classified according to their phenotype based on the International Sepsis Forum Consensus Conference definition (Calandra & Cohen, 2005) and on a *post hoc* review of all available clinical, radiologic, and microbiologic evidence. For that study, the bulk RNA extracted from peripheral blood cells and the Human Genome U219 96-array (Thermo-Fisher Scientific) were used to obtain the gene expression profiles. Based on that, gene expression comparisons for this study were based on 108 patients with CAP and 42 healthy controls.

**Supplementary Results**

**Figure S1. Regional plot of the association results for the chromosomes 4q28.2, 6p21.32, 11p12, and 20q11.22.** The y-axis displays the transformed *p*-values (-log_10_ *p*-value) while the x-axis represents chromosome positions (GRCh37/hg19) with the approximate locations of the genes encoded in the region. The horizontal dashed line represents the genome-wide significance threshold (*p*=5.0x10^-8^). Linkage disequilibrium (r^2^) values are based on the European population and is represented according to the color scheme of the top left inset. The plots were generated with LocusZoom (<http://locuszoom.org/>).

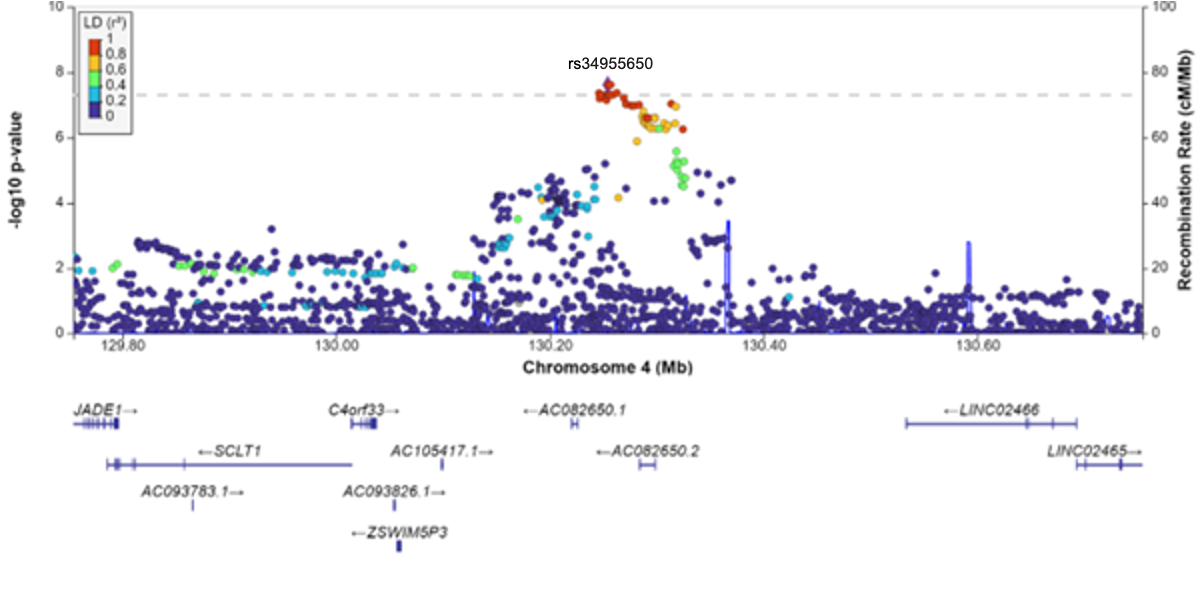

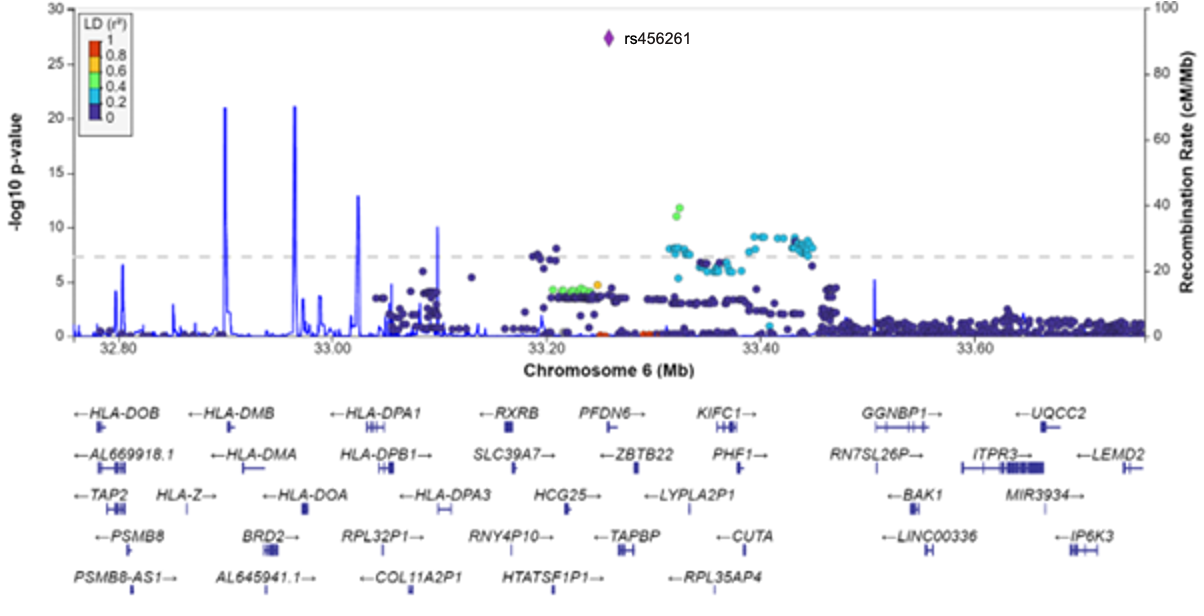

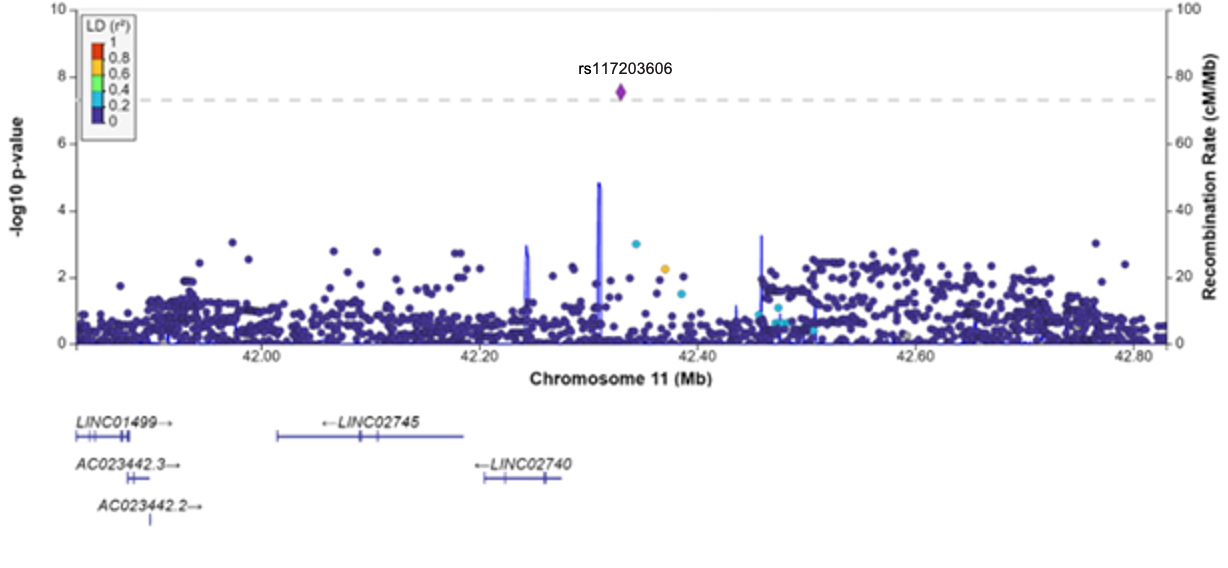

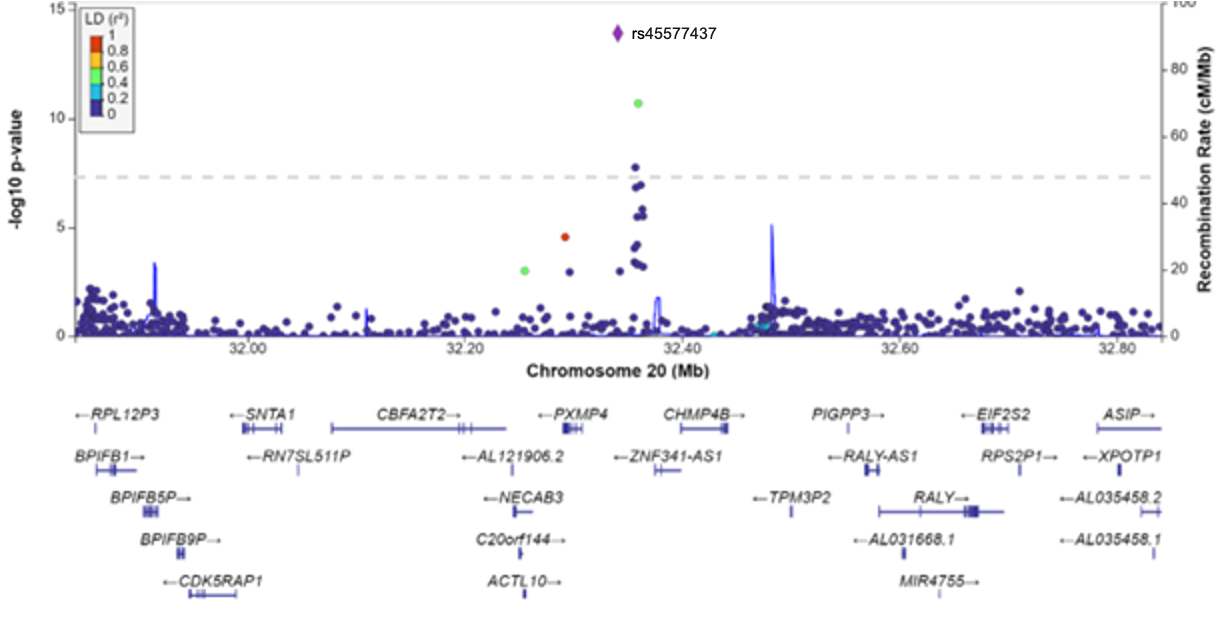

**Figure S2. Results of the Bayesian fine mapping for chromosomes 4q28.2 and 6p21.32.** The y-axis displays the transformed *p*-values (-log_10_ P-value) while the x-axis represents chromosome positions (GRCh37/hg19). The orange diamond represents the most significant SNP of each region and the variants included in the credible sets are highlighted in green.

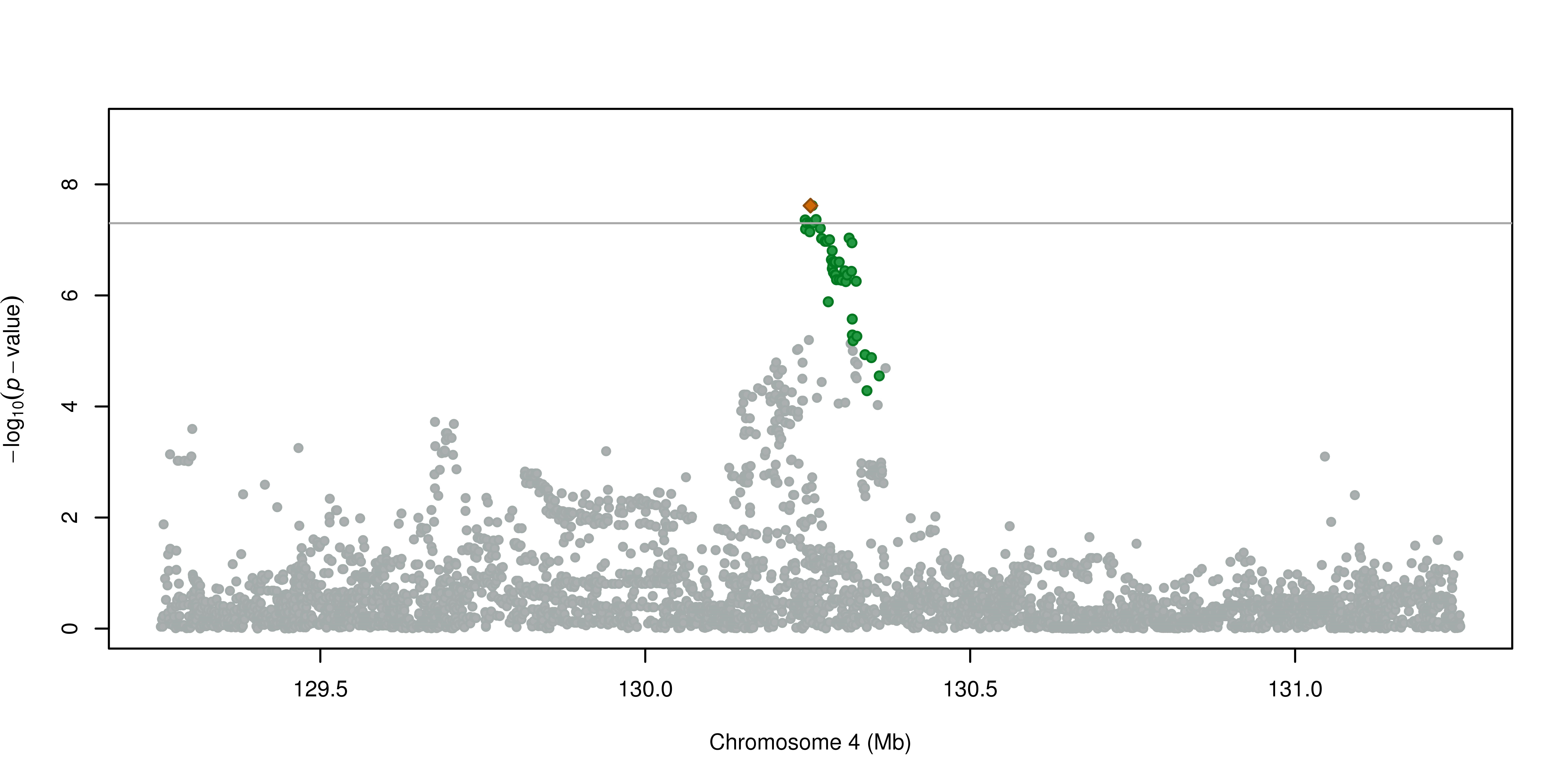

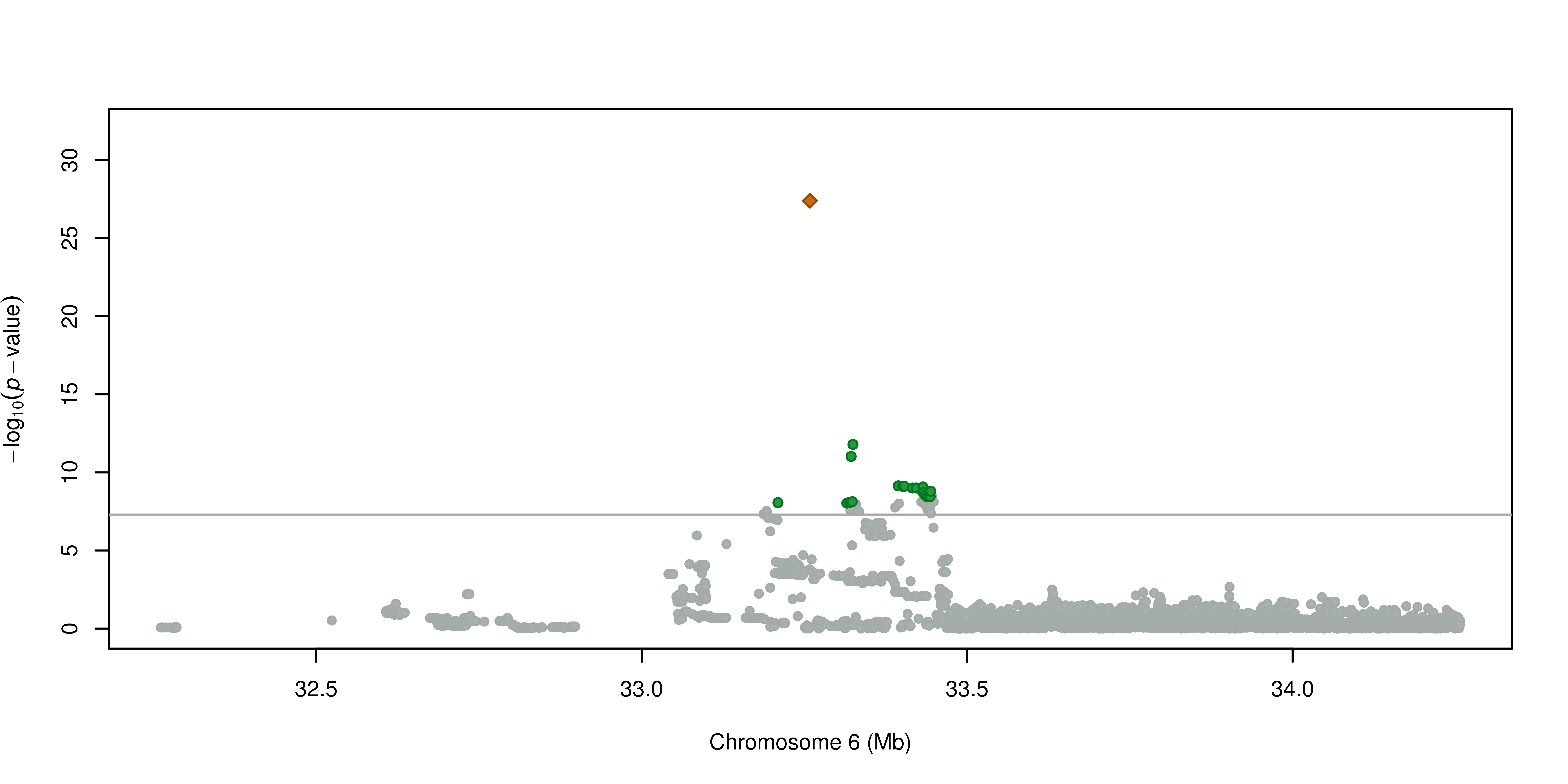

**Figure S3. Gene expression differences between ICU patients with CAP and controls for *TABPB, C4orf33,* and *ZNF341.*** Probe intensities of expression arrays were obtained from blood samples from 42 healthy controls and 108 ICU patients with CAP. Differences in average intensities were statistically assessed using t-test. The probes used for each of the genes are indicated in parentheses in the title of each comparison. Data was obtained from the GEO accession GSE65682.

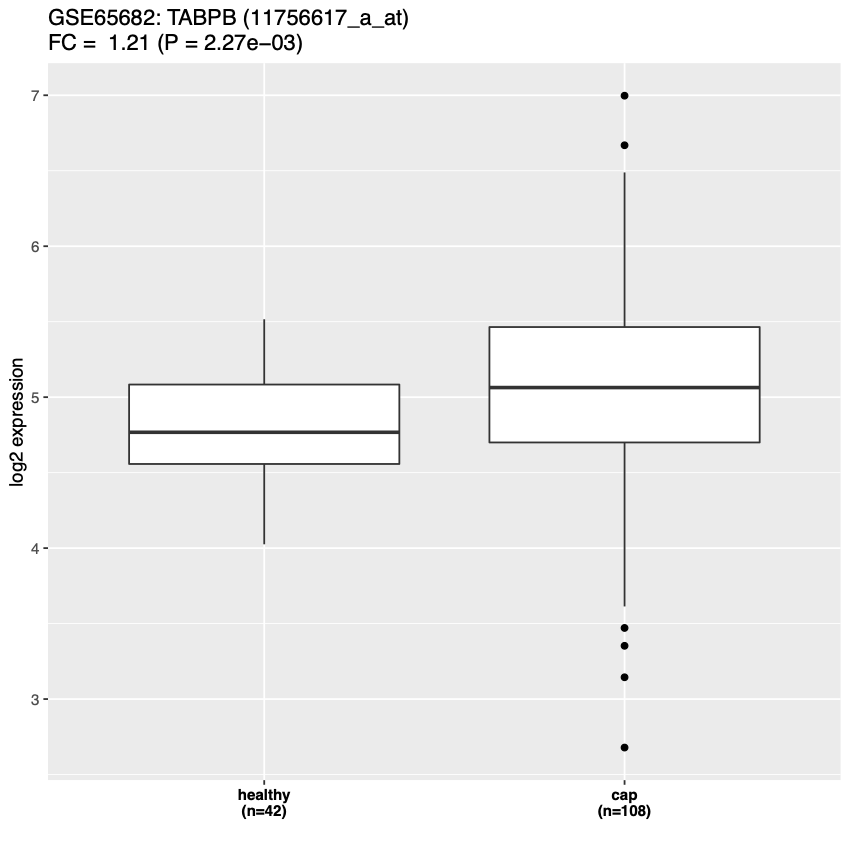

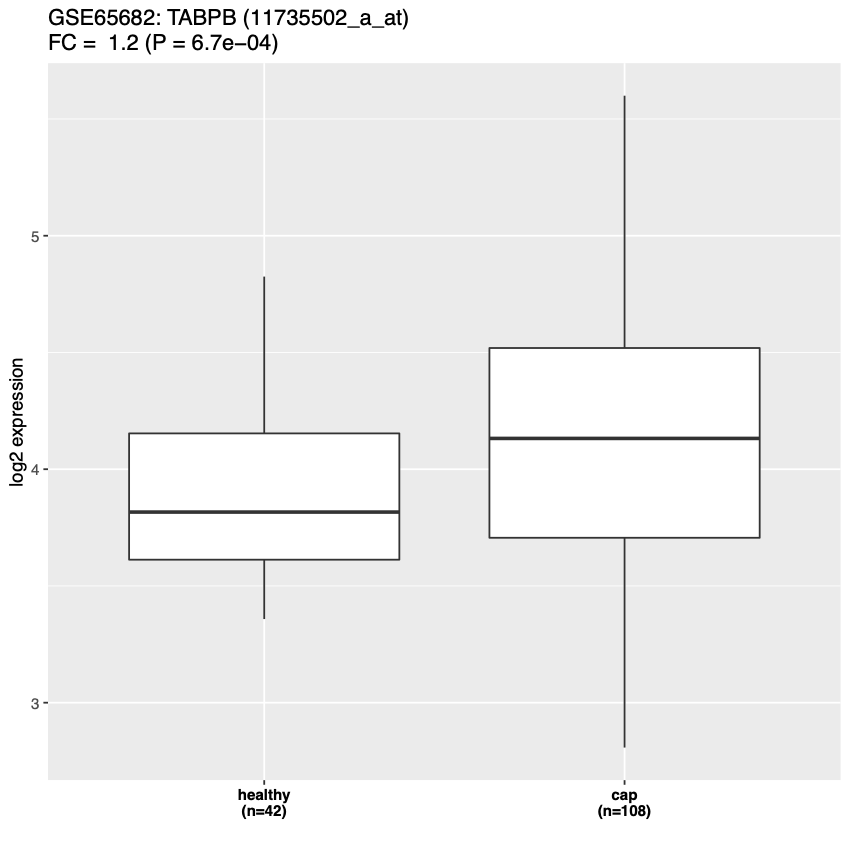

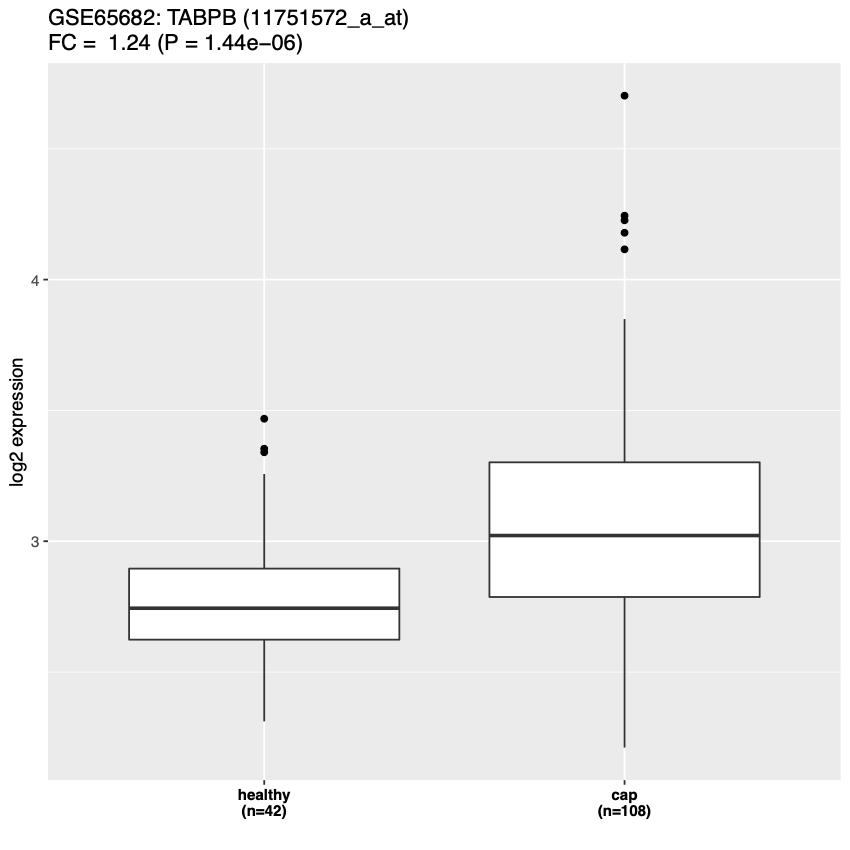

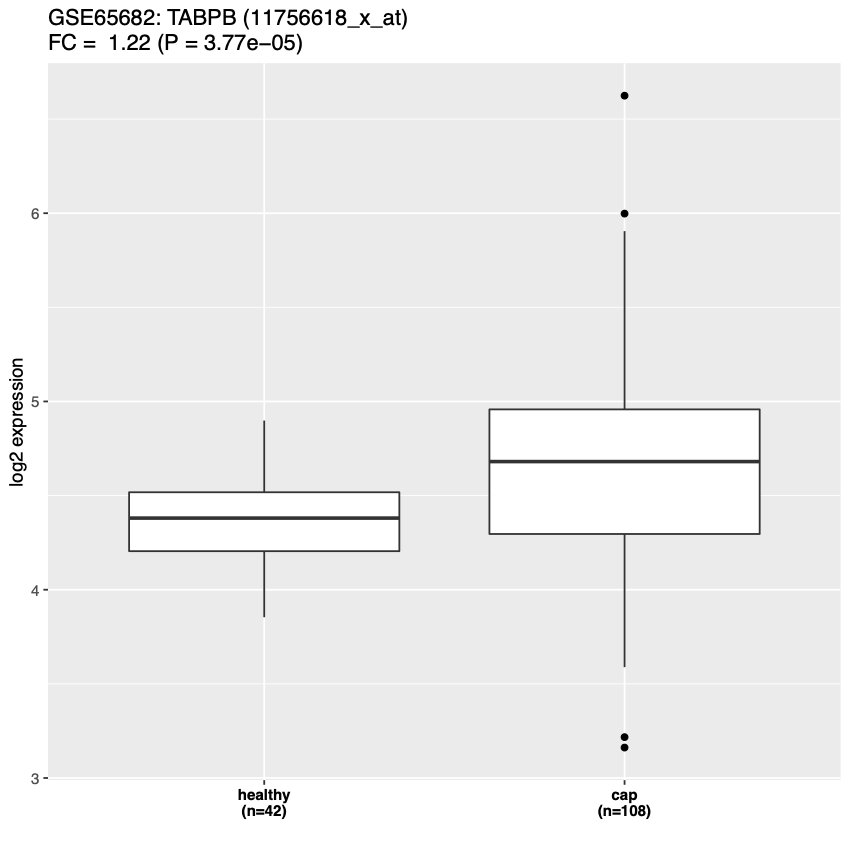

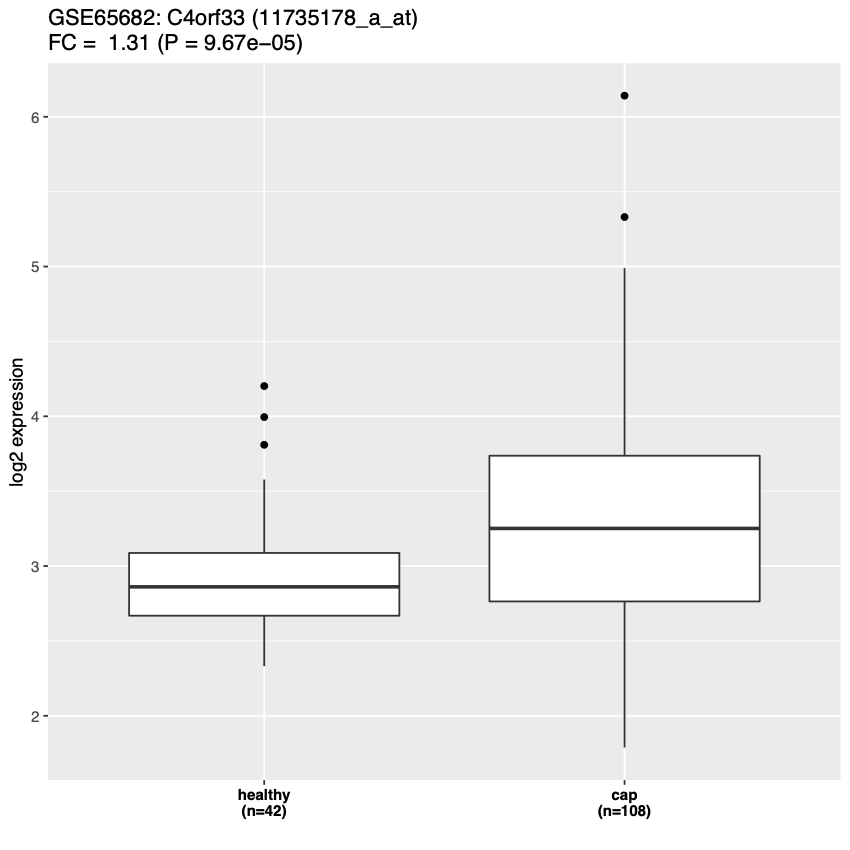

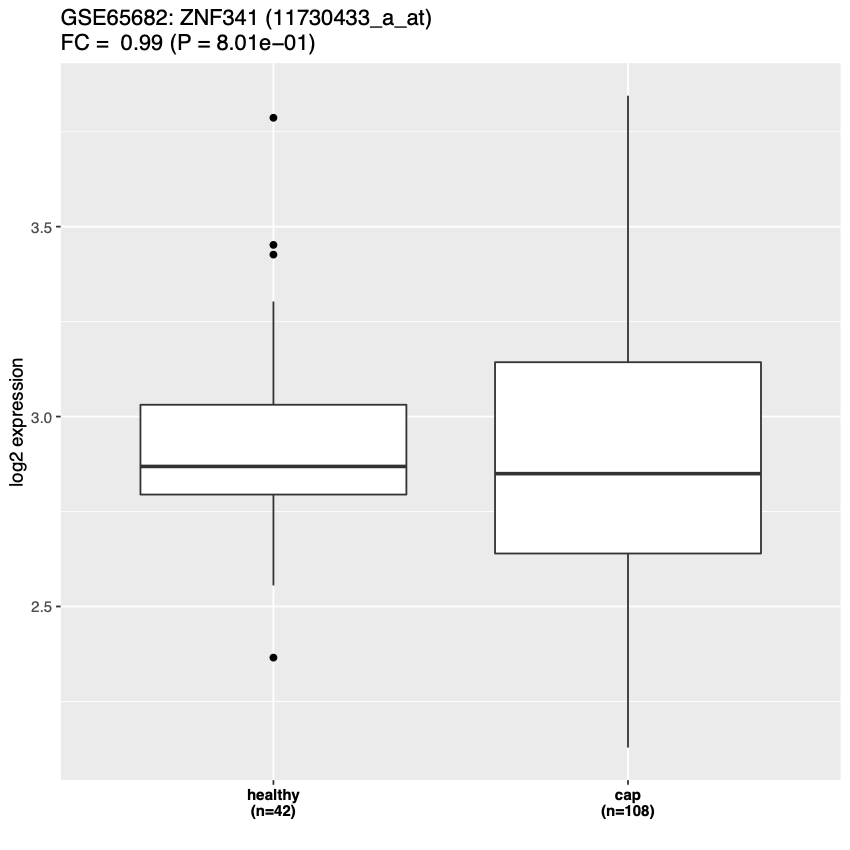

**Figure S4. Association results of the HLA region for the 155 classical HLA alleles and the 2584 amino acids for the eight *HLA* genes.** The y-axis displays the transformed *p*-values (-log_10_ *p*-values) while the x-axis represents chromosome positions (GRCh37/hg19). The significance threshold was established with a Bonferroni correction based on the number of different alleles (at *p*<4.35x10^-4^ for the HLA classical alleles and *p*<1.93x10^-5^ for the amino acids). For comparison, the association results from the SNPs included in the GWAS are represented in grey.

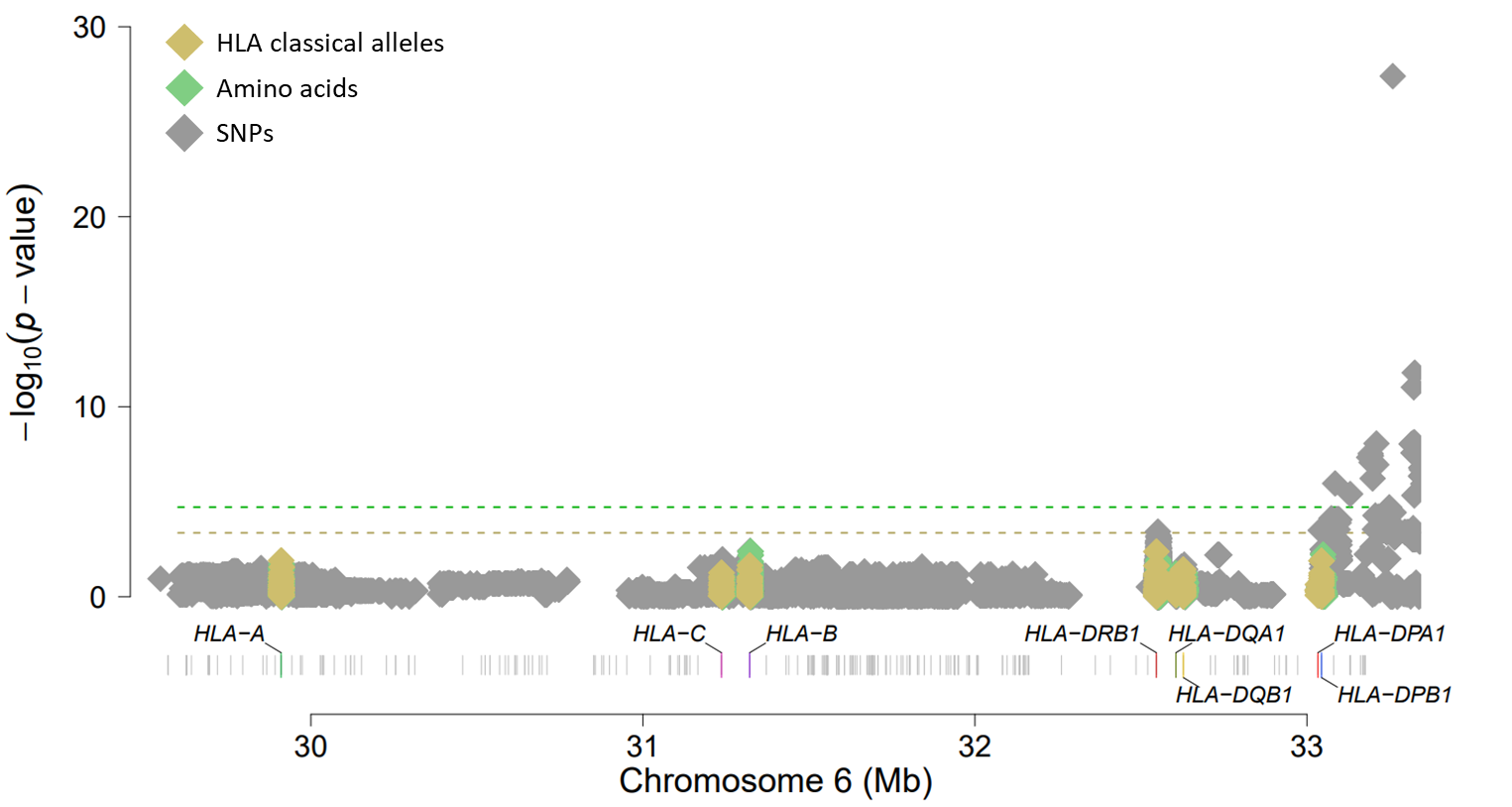

| **Table S1. Clinical and demographic characteristics of the subjects used for the genome-wide association study.** | | | |
| --- | --- | --- | --- |
|  | Cases (n=257) | Controls (n=3,508) | *p-value** |
| Age, mean years (± SD) | 64.25 ± 17.65 | 48.09 ± 14.28 | 2.2 x 10^-6^ |
| Male, count (%) | 165/257 (64.20%) | 1,911/3,508 (54.47%) | 3.1 x 10^-3^ |
| Smokers: |  |  |  |
| Non-Smokers | 106/255 (41.57%) | NA |  |
| Ever-smokers | 113/255 (44.31%) | NA |  |
| Ex-Smokers ** | 36/255 (14.12%) | NA |  |
| Comorbidities: |  |  |  |
| No comorbidities | 64/257 (24.90%) | NA |  |
| COPD | 74/255 (28.79%) | NA |  |
| Asthma | 10/257 (3.89%) | NA |  |
| Neoplasia | 30/257 (11.67%) | NA |  |
| Ischemic cardiopathy | 47/257 (18.29%) | NA |  |
| Diabetes | 67/257 (26.07%) | NA |  |
| Renal insufficiency | 34/257 (13.23%) | NA |  |
| Hepatic insufficiency | 9/257 (3.50%) | NA |  |
| Neurologic disease | 42/256 (16.34%) | NA |  |
| Autoimmune disease | 10/257 (3.89%) | NA |  |
| Psychiatric disease | 17/257 (6.61%) | NA |  |
| Clinical features: |  |  |  |
| Severe sepsis | 21/232 (9.05%) | NA |  |
| Septic shock | 38/232 (16.38%) | NA |  |
| Severe respiratory insufficiency | 161/256 (62.89%) | NA |  |
| ARDS | 8/229 (3.49%) | NA |  |
| MOF | 33/224 (14.73%) | NA |  |
| Bacteremia | 25/257 (9.73%) | NA |  |
| Pleural effusion | 44/240 (18.33%) | NA |  |
| 90-day mortality | 15/256 (5.86%) | NA |  |
| Pneumonia severity index |  |  |  |
| I-III (low) | 123/249 (49.40%) |  |  |
| IV-V (moderate-high) | 126/249 (50.60%) |  |  |
| Etiology |  |  |  |
| Non-definite etiology | 180/257 (70.04%) |  |  |
| *S. pneumoniae^~~#~~^* | 77/257 (29.96%) |  |  |
| All participants have age, and sex information. For the rest of recorded variables, percentages are accompanied by the counts out from the patients with non-missing information. NA: not available; COPD: chronic obstructive pulmonary disease; ARDS: acute respiratory distress syndrome; MOF: multiple-organ failure. *Statistical significances were obtained using the Wilcoxon test (age) and the Chi-squared test (sex). ^#^Including four patients with coinfection by *Moraxella catharralis* (1 patient), *Candida* spp. (2 patients) and influenza and parainfluenza virus (1 patient). | | | |

| Table S2. Variants that were genome-wide significant (*p*<5.0x10^-8^). | | | | | | | |
| --- | --- | --- | --- | --- | --- | --- | --- |
| rsID | Chr:Position | *p*-value | MAF | A1/A2 | OR[95% CI] | Func.Gene | Nearby gene(s) |
| rs34864289 | 4:130246086 | 4.36x10^-8^ | 0.05 | G/A | 2.45[1.78-3.37] | intergenic | *C4orf33\LINC02466* |
| rs35796550 | 4:130248440 | 4.93x10^-8^ | 0.05 | C/T | 2.43[1.76-3.34] | intergenic | *C4orf33\LINC02466* |
| rs34974459 | 4:130253077 | 4.93x10^-8^ | 0.05 | A/G | 2.43[1.76-3.34] | intergenic | *C4orf33\LINC02466* |
| rs34955650 | 4:130254264 | 2.41x10^-8^ | 0.05 | C/T | 2.46[1.80-3.38] | intergenic | *C4orf33\LINC02466* |
| rs17014599 | 4:130254717 | 4.93x10^-8^ | 0.05 | A/G | 2.43[1.76-3.34] | intergenic | *C4orf33\LINC02466* |
| rs10518546 | 4:130254808 | 2.41x10^-8^ | 0.05 | G/A | 2.46[1.18-3.38] | intergenic | *C4orf33\LINC02466* |
| rs17014611 | 4:130255450 | 4.93x10^-8^ | 0.05 | G/A | 2.43[1.76-3.34] | intergenic | *C4orf33\LINC02466* |
| rs13109938 | 4:130256289 | 4.93x10^-8^ | 0.05 | C/A | 2.43[1.76-3.34] | intergenic | *C4orf33\LINC02466* |
| rs35004602 | 4:130256929 | 2.41x10^-8^ | 0.05 | G/C | 2.46[1.80-3.38] | intergenic | *C4orf33\LINC02466* |
| rs35255990 | 4:130258899 | 4.93x10^-8^ | 0.05 | C/T | 2.43[1.76-3.34] | intergenic | *C4orf33\LINC02466* |
| rs17367613 | 4:130262824 | 4.30x10^-8^ | 0.05 | G/C | 2.44[1.77-3.35] | intergenic | *C4orf33\LINC02466* |
| rs213211 | 6:33187355 | 4.64x10^-8^ | 0.03 | C/T | 2.64[1.86-3.74] | intergenic | *RING1\HCG25* |
| rs437119 | 6:33191784 | 2.94x10^-8^ | 0.04 | T/C | 2.60[1.86-3.65] | intergenic | *RING1\HCG25* |
| rs439121 | 6:33192867 | 3.87x10^-8^ | 0.03 | A/C | 2.66[1.87-3.76] | intergenic | *RING1\HCG25* |
| rs213226 | 6:33209310 | 8.64x10^-9^ | 0.17 | A/G | 1.84[1.49-2.26] | intergenic | *RING1\HCG25* |
| rs456261 | 6:33258443 | 4.00x10^-28^ | 0.02 | G/A | 8.23[5.65-11.99] | intronic | *PFDN6* |
| rs3130272 | 6:33315086 | 9.19x10^-9^ | 0.16 | G/C | 0.54[0.44-0.66] | intergenic | *SMIM40\KIFC1* |
| rs211476 | 6:33319596 | 8.11x10^-9^ | 0.16 | G/T | 0.54[0.43-0.66] | intergenic | *SMIM40\KIFC1* |
| rs211475 | 6:33320089 | 8.11x10^-9^ | 0.16 | A/G | 0.54[0.43-0.66] | intergenic | *SMIM40\KIFC1* |
| rs211474 | 6:33320613 | 2.71x10^-8^ | 0.16 | G/A | 0.55[0.44-0.68] | intergenic | *SMIM40\KIFC1* |
| rs211473 | 6:33320759 | 8.11x10^-9^ | 0.16 | G/A | 0.54[0.43-0.66] | intergenic | *SMIM40\KIFC1* |
| rs3130008 | 6:33321666 | 9.48x10^-12^ | 0.08 | G/A | 2.45[1.89-3.17] | intergenic | *SMIM40\KIFC1* |
| rs211467 | 6:33323446 | 7.53x10^-9^ | 0.16 | G/C | 0.54[0.43-0.66] | intergenic | *SMIM40\KIFC1* |
| rs3117324 | 6:33324527 | 1.62x10^-12^ | 0.08 | G/T | 2.51[1.95-3.24] | intergenic | *SMIM40\KIFC1* |
| rs211455 | 6:33328518 | 1.03x10^-8^ | 0.16 | G/A | 0.54[0.44-0.67] | intergenic | *SMIM40\KIFC1* |
| rs211454 | 6:33329068 | 1.03x10^-8^ | 0.16 | G/A | 0.54[0.44-0.67] | intergenic | *SMIM40\KIFC1* |
| rs211453 | 6:33330131 | 2,82x10^-8^ | 0.16 | T/G | 0.55[0.44-0.68] | intergenic | *SMIM40\KIFC1* |
| rs211452 | 6:33330981 | 2,82x10^-8^ | 0.16 | A/G | 0.55[0.44-0.68] | intergenic | *SMIM40\KIFC1* |
| rs211450 | 6:33333446 | 2.93x10^-8^ | 0.16 | C/T | 0.55[0.44-0.68] | intergenic | *SMIM40\KIFC1* |
| rs211449 | 6:33333916 | 3.11x10^-8^ | 0.16 | G/T | 0.55[0.44-0.68] | intergenic | *SMIM40\KIFC1* |
| rs211456 | 6:33389381 | 1.77x10^-8^ | 0.16 | G/T | 0.54[0.44-0.67] | intronic | *SYNGAP1* |
| rs2076775 | 6:33394253 | 7,26x10^-10^ | 0.14 | C/G | 0.50[0.41-0.63] | intronic | *SYNGAP1* |
| rs9461856 | 6:33395199 | 9.90x10^-9^ | 0.08 | G/A | 2.16[1.66-2.82] | intronic | *SYNGAP1* |
| rs381847 | 6:33401124 | 7.69x10^-10^ | 0.14 | C/G | 0.50[0.41-0.63] | intronic | *SYNGAP1* |
| rs453590 | 6:33403422 | 7.69x10^-10^ | 0.14 | C/T | 0.50[0.41-0.63] | intronic | *SYNGAP1* |
| rs2772376 | 6:33416111 | 9.94x10^-10^ | 0.14 | C/T | 0.51[0.41-0.63] | intronic | *SYNGAP1* |
| rs2247385 | 6:33421577 | 9.94x10^-10^ | 0.14 | T/C | 0.51[0.41-0.63] | upstream\  downstream | *ZBTB9\SYNGAP1* |
| rs2772373 | 6:33429672 | 7.64x10^-9^ | 0.03 | A/G | 2.90[2.02-4.17] | intergenic | *ZBTB9\BAK1* |
| rs1613451 | 6:33430701 | 7.64x10^-9^ | 0.03 | T/C | 2.90[2.02-4.17] | intergenic | *ZBTB9\BAK1* |
| rs1755047 | 6:33431319 | 7.64x10^-9^ | 0.03 | G/C | 2.90[2.02-4.17] | intergenic | *ZBTB9\BAK1* |
| rs2772375 | 6:33431569 | 1.78x10^-9^ | 0.14 | T/C | 0.51[0.41-0.63] | intergenic | *ZBTB9\BAK1* |
| rs1755048 | 6:33432160 | 8.46x10^-10^ | 0.14 | G/A | 0.50[0.41-0.63] | intergenic | *ZBTB9\BAK1* |
| rs4711325 | 6:33432709 | 1,96x10^-9^ | 0.21 | A/G | 0.54[0.44-0.66] | intergenic | *ZBTB9\BAK1* |
| rs2772389 | 6:33435179 | 2.87x10^-9^ | 0.14 | G/A | 0.52[0.42-0.64] | intergenic | *ZBTB9\BAK1* |
| rs2772390 | 6:33436144 | 1.29x10^-8^ | 0.03 | A/G | 2.85[1.99-4.09] | intergenic | *ZBTB9\BAK1* |
| rs1755049 | 6:33438346 | 3.53x10^-9^ | 0.14 | C/T | 0.52[0.41-0.64] | intergenic | *ZBTB9\BAK1* |
| rs1755046 | 6:33438359 | 3.53x10^-9^ | 0.14 | G/A | 0.52[0.41-0.64] | intergenic | *ZBTB9\BAK1* |
| rs2772391 | 6:33438487 | 3.53x10^-9^ | 0.14 | C/T | 0.52[0.41-0.64] | intergenic | *ZBTB9\BAK1* |
| rs4713625 | 6:33438509 | 3.53x10^-9^ | 0.14 | T/C | 0.52[0.41-0.64] | intergenic | *ZBTB9\BAK1* |
| rs2772392 | 6:33438883 | 2.34x10^-8^ | 0.03 | T/C | 2.79[1.95-4.01] | intergenic | *ZBTB9\BAK1* |
| rs9283903 | 6:33439777 | 3.53x10^-9^ | 0.14 | C/T | 0.52[0.41-0.64] | intergenic | *ZBTB9\BAK1* |
| rs13198907 | 6:33441648 | 6.11x10^-9^ | 0.08 | T/A | 2.16[1.67-2.81] | intergenic | *ZBTB9\BAK1* |
| rs3130034 | 6:33441993 | 2.34x10^-8^ | 0.03 | A/G | 2.79[1.95-4.01] | intergenic | *ZBTB9\BAK1* |
| rs3130031 | 6:33442384 | 2.34x10^-8^ | 0.03 | A/C | 2.79[1.95-4.01] | intergenic | *ZBTB9\BAK1* |
| rs3130030 | 6:33442652 | 2.34x10^-8^ | 0.03 | T/C | 2.79[1.95-4.01] | intergenic | *ZBTB9\BAK1* |
| rs3130028 | 6:33442738 | 2.34x10^-8^ | 0.03 | C/T | 2.79[1.95-4.01] | intergenic | *ZBTB9\BAK1* |
| rs62407928 | 6:33442816 | 2.34x10^-8^ | 0.03 | T/C | 2.79[1.95-4.01] | intergenic | *ZBTB9\BAK1* |
| rs2772393 | 6:33443189 | 3.53x10^-9^ | 0.14 | G/A | 0.52[0.41-0.64] | intergenic | *ZBTB9\BAK1* |
| rs2772394 | 6:33443907 | 1.07x10^-8^ | 0.03 | G/A | 2.87[2.00-4.12] | intergenic | *ZBTB9\BAK1* |
| rs9366821 | 6:33444074 | 1.61x10^-9^ | 0.14 | T/G | 0.51[0.41-0.64] | intergenic | *ZBTB9\BAK1* |
| rs3119023 | 6:33444395 | 4.27x10^-8^ | 0.03 | A/C | 2.74[1.91-3.92] | intergenic | *ZBTB9\BAK1* |
| rs3130025 | 6:33445962 | 3.47x10^-9^ | 0.03 | G/T | 2.91[2.04-4.15] | intergenic | *ZBTB9\BAK1* |
| rs2480487 | 6:33448613 | 7.71x10^-9^ | 0.03 | C/A | 2.84[1.99-4.04] | intergenic | *ZBTB9\BAK1* |
| rs117203606 | 11:42330132 | 2.90x10^-8^ | 0.11 | G/A | 0.21[0.12-0.36] | intergenic | *LINC02740\*  *HNRNPKP3* |
| rs45577437 | 20:32341041 | 1.21x10^-14^ | 0.44 | C/T | 0.46[0.37-0.56] | exonic | *ZNF341* |
| rs2747557 | 20:32357157 | 1.77x10^-8^ | 0.46 | A/G | 0.56[0.45-0.68] | intronic | *ZNF341* |
| rs115879816 | 20:32359799 | 2.02x10^-11^ | 0.29 | G/A | 0.41[0.32-0.54] | intronic | *ZNF341* |
| MAF: Minor allele frequency; A1: Non-effect allele; A2: Effect allele; OR: odd ratio; CI: confidence interval; Func.Gene: predicted function. | | | | | | | |

| Table S3. Results of the sentinel variants that were independently associated with CAP in models adjusting for age, sex, and the three main principal components of genetic heterogeneity. | | | |
| --- | --- | --- | --- |
| rsID | Position | OR[95% CI] | *p-*value |
| rs34955650 | 4:130254264 | 2.51[1.80-3.50] | 1.24x10^-7^ |
| rs213226 | 6:33209310 | 1.80[1.43-2.28] | 3.74x10^-7^ |
| rs456261 | 6:33258443 | 7.17[4.66-11.04] | 5.87x10^-19^ |
| rs2076775 | 6:33394253 | 0.50[0.40-0.63] | 8.47x10^-9^ |
| rs117203606 | 11:42330132 | 0.21[0.12-0.36] | 7.87x10^-8^ |
| rs45577437 | 20:32341041 | 0.43[0.34-0.53] | 8.59x10^-15^ |
| Position: chromosome and base pair according to GRCh37/hg19; OR: odds ratio; CI: confidence interval. | | | |

| Table S4. Results of the sentinel variants independently associated with severe outcomes. Models were adjusted for sex and the three main principal components of genetic heterogeneity. | | | | | | | |
| --- | --- | --- | --- | --- | --- | --- | --- |
| Genetic variant | | SS/SSh and SRI | | SS/SSh | | SRI | |
| rsID | Position | *p*-value | OR[95% CI] | *p*-value | OR[95% CI] | *p*-value | OR[95% CI] |
| rs34955650 | 4:130254264 | 3.54x10^-4^ | 3.07[1.66-5.69] | 7.29x10^-5^ | 3.21[1.80-5.70] | 3.24x10^-6^ | 2.50[1.70-3.68] |
| rs213226 | 6:33209310 | 0.02 | 1.74[1.09-2.79] | 0.03 | 1.66[1.06-2.60] | 1.02x10^-5^ | 1.79[1.38-2.32] |
| rs456261 | 6:33258443 | 3.29x10^-9^ | 8.66[4.24-17.72] | 7.23x10^-10^ | 8.50[4.30-16.78] | 2.13x10^-22^ | 8.87[5.72-13.77] |
| rs2076775 | 6:33394253 | 4.33x10^-3^ | 0.49[0.30-0.80] | 8.37x10^-4^ | 0.46[0.29-0.73] | 4.51x10^-7^ | 0.50[0.38-0.65] |
| rs117203606 | 11:42330132 | 0.02 | 0.24[0.07-0.77] | 8.67x10^-3^ | 0.21[0.06-0.67] | 1.48x10^-5^ | 0.23[0.12-0.44] |
| rs45577437 | 20:32341041 | 1.13x10^-4^ | 0.38[0.23-0.62] | 2.13x10^-5^ | 0.37[0.23-0.58] | 8.92x10^-11^ | 0.43[0.33-0.55] |
| All the association tests were assessed by considering the 3,508 controls and one of the following groups of CAP patients as cases: i) 52 patients with both severe respiratory insufficiency and severe sepsis or septic shock; ii) 58 patients with severe sepsis or septic shock; and iii) 161 patients with severe respiratory insufficiency. Position: chromosome and base pair according to GRCh37/hg19; OR: odds ratio; CI: confidence interval. SS/SSh: severe sepsis or septic shock. SRI: severe respiratory insufficiency. | | | | | | | |

| Table S5. Association results for the variants included in the credible sets of the Bayesian fine mapping of 4q28.2 and 6p21.32 | | | | | | | | | |
| --- | --- | --- | --- | --- | --- | --- | --- | --- | --- |
| rsID | Position_A1/A2 | MAF | p-value | OR[95%CI] | Biotype [VEP] | CADD score | Gene  [V2G] | Best score  [V2G] | Consequence |
| rs34864289 | 4:130246086_G/A | 0.05 | 4.36x10^-8^ | 2.45[1.78-3.37] | promoter flanking region | 3.43 | *C4orf33* | 0.060 | low |
| rs11943511 | 4:130246680_C/T | 0.05 | 6.37x10^-8^ | 2.41[1.75-3.31] | promoter flanking region | 0.66 | *C4orf33* | 0.060 | low |
| rs35796550 | 4:130248440_C/T | 0.05 | 4.93x10^-8^ | 2.43[1.76-3.34] | promoter flanking region | 1.41 | *C4orf33* | 0.086 | low |
| rs34974459 | 4:130253077_A/G | 0.05 | 4.93x10^-8^ | 2.43[1.76-3.34] | - | 1.06 | *C4orf33* | 0.073 | low |
| rs34466472 | 4:130253129_G/A | 0.05 | 7.14x10^-8^ | 2.39[1.74-3.28] | - | 4.47 | *C4orf33* | 0.073 | Low |
| rs34955650 | 4:130254264_C/T | 0.05 | 2.41x10^-8^ | 2.46[1.80-3.38] | - | 7.75 | *C4orf33* | 0.060 | High |
| rs17014599 | 4:130254717_A/G | 0.05 | 4.93x10^-8^ | 2.43[1.76-3.34] | - | 1.77 | *C4orf33* | 0.066 | Low |
| rs10518546 | 4:130254808_G/A | 0.05 | 2.41x10^-8^ | 2.46[1.80-3.38] | - | 2.14 | *C4orf33* | 0.066 | Low |
| rs17014611 | 4:130255450_G/A | 0.05 | 4.93x10^-8^ | 2.43[1.76-3.34] | - | 18.23 | *C4orf33* | 0.060 | High |
| rs13109938 | 4:130256289_C/A | 0.05 | 4.93x10^-8^ | 2.43[1.76-3.34] | - | 19.34 | *C4orf33* | 0.060 | High |
| rs35004602 | 4:130256929_G/C | 0.05 | 2.41x10^-8^ | 2.46[1.80-3.38] | enhancer | 15.70 | *C4orf33* | 0.060 | High |
| rs35255990 | 4:130258899_C/T | 0.05 | 4.93x10^-8^ | 2.43[1.76-3.34] | - | 0.47 | *C4orf33* | 0.060 | Low |
| rs17367613 | 4:130262824_G/C | 0.05 | 4.30x10^-8^ | 2.44[1.77-3.35] | - | 1.52 | *C4orf33* | 0.086 | Low |
| rs17014614 | 4:130269324_A/G | 0.05 | 6.15x10^-8^ | 2.44[1.76-3.36] | - | 11.03 | *C4orf33* | 0.060 | High |
| rs35918555 | 4:130271319_G/A | 0.05 | 9.44x10^-8^ | 2.40[1.74-3.32] | - | 1.19 | *C4orf33* | 0.060 | Low |
| rs13130195 | 4:130271741_C/A | 0.05 | 9.44x10^-8^ | 2.40[1.74-3.32] | - | 0.77 | *C4orf33* | 0.060 | Low |
| rs13128188 | 4:130276663_G/T | 0.05 | 1.06x10^-7^ | 2.40[1.74-3.31] | enhancer | 11.87 | *C4orf33* | 0.060 | High |
| rs35524133 | 4:130277071_G/T | 0.05 | 1.06x10^-7^ | 2.40[1.74-3.31] | enhancer | 9.10 | *C4orf33* | 0.060 | High |
| rs13149187 | 4:130279342_G/A | 0.05 | 1.06x10^-7^ | 2.40[1.74-3.31] | - | 0.33 | *C4orf33* | 0.060 | Low |
| rs28580176 | 4:130281632_C/T | 0.06 | 1.30x10^-6^ | 2.12[1.56-2.87] | - | 8.26 | *C4orf33* | 0.060 | High |
| rs13102645 | 4:130283572_T/G | 0.05 | 9.93x10^-8^ | 2.39[1.73-3.29] | CTCF binding site | 1.97 | *C4orf33* | 0.060 | Low |
| rs71613927 | 4:130286444_C/T | 0.05 | 2.27x10^-7^ | 2.30[1.68-3.16] | CTCF binding site | 1.93 | *C4orf33* | 0.060 | Low |
| rs79136817 | 4:130287632_C/A | 0.05 | 1.56x10^-7^ | 2.32[1.69-3.17] | - | 0.36 | *C4orf33* | 0.046 | Low |
| rs71613928 | 4:130287805_A/G | 0.05 | 3.28x10^-7^ | 2.25[1.65-3.07] | - | 0.54 | *C4orf33* | 0.060 | Low |
| rs34099241 | 4:130288481_A/G | 0.05 | 2.51x10^-7^ | 2.30[1.67-3.15] | - | 3.17 | *C4orf33* | 0.060 | Low |
| rs11942914 | 4:130288620_A/G | 0.05 | 2.97x10^-7^ | 2.26[1.65-3.08] | - | 5.49 | *C4orf33* | 0.060 | High |
| rs34714171 | 4:130289596_G/A | 0.05 | 3.87x10^-7^ | 2.24[1.64-3.06] | - | 1.59 | *C4orf33* | 0.053 | Low |
| rs34010234 | 4:130290261_C/T | 0.05 | 2.50x10^-7^ | 2.31[1.68-3.18] | - | 2.32 | *C4orf33* | 0.053 | Low |
| rs13152421 | 4:130291896_C/G | 0.05 | 4.29x10^-7^ | 2.24[1.64-3.07] | - | 14.26 | *C4orf33* | 0.053 | High |
| rs71613929 | 4:130292532_C/A | 0.05 | 2.50x10^-7^ | 2.31[1.68-3.18] | - | 0.15 | *C4orf33* | 0.053 | Low |
| rs11940099 | 4:130293025_G/C | 0.05 | 4.29x10^-7^ | 2.24[1.64-3.07] | - | 2.02 | *C4orf33* | 0.053 | Low |
| rs71613930 | 4:130294285_T/C | 0.05 | 5.21x10^-7^ | 2.23[1.63-3.05] | - | 0.24 | *C4orf33* | 0.053 | Low |
| rs71613931 | 4:130298354_A/G | 0.05 | 2.50x10^-7^ | 2.31[1.68-3.18] | enhancer | 4.68 | *C4orf33* | 0.053 | Low |
| rs71613932 | 4:130298375_A/G | 0.05 | 2.51x10^-7^ | 2.30[1.67-3.15] | enhancer | 2.57 | *C4orf33* | 0.053 | Low |
| rs35618244 | 4:130298386_T/G | 0.05 | 2.51x10^-7^ | 2.30[1.67-3.15] | enhancer | 6.06 | *C4orf33* | 0.053 | High |
| rs13135051 | 4:130298557_T/C | 0.05 | 5.21x10^-7^ | 2.23[1.63-3.05] | - | 0.85 | *C4orf33* | 0.053 | Low |
| rs11944963 | 4:130302263_C/T | 0.06 | 5.33x10^-7^ | 2.13[1.58-2.86] | - | 3.92 | *C4orf33* | 0.040 | Low |
| rs36062368 | 4:130307035_C/G | 0.05 | 3.61x10^-7^ | 2.26[1.65-3.09] | - | 5.78 | *C4orf33* | 0.053 | High |
| rs4975219 | 4:130308675_T/C | 0.05 | 5.63x10^-7^ | 2.22[1.63-3.04] | - | 4.40 | *C4orf33* | 0.040 | Low |
| rs13120481 | 4:130310638_A/G | 0.05 | 4.28x10^-7^ | 2.24[1.64-3.07] | - | 0.30 | *C4orf33* | 0.053 | Low |
| rs71613934 | 4:130313713_G/A | 0.05 | 9.27x10^-8^ | 2.37[1.73-3.25] | - | 0.17 | *C4orf33* | 0.053 | Low |
| rs6845051 | 4:130317282_T/G | 0.05 | 3.69x10^-7^ | 2.24[1.64-3.06] | - | 2.49 | *C4orf33* | 0.040 | Low |
| rs13139447 | 4:130318033_G/T | 0.05 | 1.13x10^-7^ | 2.34[1.71-3.20] | - | 1.00 | *C4orf33* | 0.053 | Low |
| rs13115941 | 4:130318517_T/C | 0.06 | 2.66x10^-6^ | 2.00[1.50-2.66] | - | 4.63 | *C4orf33* | 0.027 | Low |
| rs13140408 | 4:130318692_A/T | 0.07 | 5.16x10^-6^ | 1.93[1.45-2.56] | - | 1.09 | *C4orf33* | 0.027 | Low |
| rs2117058 | 4:130319851_A/T | 0.07 | 6.53x10^-6^ | 1.92[1.44-2.54] | - | 0.03 | *C4orf33* | 0.027 | Low |
| rs17014727 | 4:130324558_C/T | 0.05 | 5.56x10^-7^ | 2.26[1.64-3.11] | - | 0.46 | *C4orf33* | 0.053 | Low |
| rs9968320 | 4:130325594_T/C | 0.07 | 5.45x10^-6^ | 1.95[1.46-2.60] | - | 0.56 | *C4orf33* | 0.027 | Low |
| rs11730148 | 4:130338172_C/A | 0.15 | 1.17x10^-5^ | 1.66[1.32-2.08] | - | 1.16 | *C4orf33* | 0.146 | Low |
| rs10001340 | 4:130341127_A/C | 0.16 | 5.20x10^-5^ | 1.59[1.27-1.99] | - | 1.74 | *C4orf33* | 0.133 | Low |
| rs1865174 | 4:130348020_C/T | 0.15 | 1.32x10^-5^ | 1.65[1.32-2.07] | - | 2.25 | *C4orf33* | 0.133 | Low |
| rs13136673 | 4:130360060_G/T | 0.15 | 2.82x10^-5^ | 1.62[1.29-2.03] | - | 7.97 | *C4orf33* | 0.133 | High |
| rs213226 | 6:33209310_A/G | 0.17 | 8.64x10^-9^ | 1.84[1.49-2.26] | processed pseudogene | 7.50 | *COL11A2* | 0.234 | Low |
| rs456261 | 6:33258443_G/A | 0.02 | 4.00x10^-28^ | 8.23[5.65-11.99] | protein coding | 5.67 | *TAPBP* | 0.410 | High |
| rs3130272 | 6:33315086_G/C | 0.16 | 9.19x10^-9^ | 0.54[0.44-0.66] | enhancer | 0.92 | *TAPBP* | 0.246 | Low |
| rs211476 | 6:33319596_G/T | 0.16 | 8.11x10^-9^ | 0.54[0.43-0.66] | open chromatin region | 4.46 | *TAPBP* | 0.246 | High |
| rs211475 | 6:33320089_A/G | 0.16 | 8.11x10^-9^ | 0.54[0.43-0.66] | - | 7.84 | *TAPBP* | 0.246 | High |
| rs211473 | 6:33320759_G/A | 0.16 | 8.11x10^-9^ | 0.54[0.43-0.66] | - | 9.19 | *TAPBP* | 0.246 | High |
| rs3130008 | 6:33321666_G/A | 0.08 | 9.48x10^-12^ | 2.45[1.89-3.17] | - | 2.85 | *TAPBP* | 0.360 | Low |
| rs211467 | 6:33323446_G/C | 0.16 | 7.53x10^-9^ | 0.54[0.43-0.66] | - | 1.18 | *TAPBP* | 0.246 | Low |
| rs3117324 | 6:33324527_G/T | 0.08 | 1.62x10^-12^ | 2.51[1.95-3.24] | promoter flanking region | 1.35 | *TAPBP* | 0.406 | Low |
| rs2076775 | 6:33394253_C/G | 0.14 | 7.26x10^-10^ | 0.50[0.41-0.63] | protein coding | 1.54 | *TAPBP* | 0.279 | Low |
| rs381847 | 6:33401124_C/G | 0.14 | 7.69x10^-10^ | 0.50[0.41-0.63] | protein coding | 10.73 | *TAPBP* | 0.232 | High |
| rs453590 | 6:33403422_C/T | 0.14 | 7.69x10^-10^ | 0.50[0.41-0.63] | protein coding | 2.55 | *TAPBP* | 0.240 | Low |
| rs2772376 | 6:33416111_C/T | 0.14 | 9.94x10^-10^ | 0.51[0.41-0.63] | protein coding | 0.17 | *TAPBP* | 0.232 | Low |
| rs2247385 | 6:33421577_T/C | 0.14 | 9.94x10^-10^ | 0.51[0.41-0.63] | protein coding | 9.41 | *TAPBP* | 0.232 | High |
| rs2772375 | 6:33431569_T/C | 0.14 | 1.78x10^-9^ | 0.51[0.41-0.63] | - | 2.26 | *TAPBP* | 0.206 | Low |
| rs1755048 | 6:33432160_G/A | 0.14 | 8.46x10^-10^ | 0.50[0.41-0.63] | - | 5.04 | *TAPBP* | 0.206 | High |
| rs4711325 | 6:33432709_A/G | 0.21 | 1.96x10^-9^ | 0.54[0.44-0.66] | - | 0.11 | *TAPBP* | 0.193 | Low |
| rs2772389 | 6:33435179_G/A | 0.14 | 2.87x10^-9^ | 0.52[0.42-0.64] | - | 0.05 | *TAPBP* | 0.206 | Low |
| rs1755049 | 6:33438346_C/T | 0.14 | 3.53x10^-9^ | 0.52[0.41-0.64] | - | 0.09 | *TAPBP* | 0.206 | Low |
| rs1755046 | 6:33438359_G/A | 0.14 | 3.53x10^-9^ | 0.52[0.41-0.64] | - | 2.37 | *TAPBP* | 0.206 | Low |
| rs2772391 | 6:33438487_C/T | 0.14 | 3.53x10^-9^ | 0.52[0.41-0.64] | - | 1.97 | *TAPBP* | 0.206 | Low |
| rs4713625 | 6:33438509_T/C | 0.14 | 3.53x10^-9^ | 0.52[0.41-0.64] | - | 3.67 | *TAPBP* | 0.206 | Low |
| rs9283903 | 6:33439777_C/T | 0.14 | 3.53x10^-9^ | 0.52[0.41-0.64] | - | 5.45 | *TAPBP* | 0.206 | High |
| rs2772393 | 6:33443189_G/A | 0.14 | 3.53x10^-9^ | 0.52[0.41-0.64] | - | 6.68 | *TAPBP* | 0.206 | High |
| rs9366821 | 6:33444074_T/G | 0.14 | 1.61x10^-9^ | 0.51[0.41-0.64] | - | 14.62 | *TAPBP* | 0.206 | High |
| Position: chromosome and base pair according to GRCh37/hg19; A1: Non-effect allele; A2: Effect allele; MAF: minor allele frequency, OR: odds ratio; CI: confidence interval; Biotype: obtained with Variant Effect Predictor (VEP) release 108; V2G: Variant-to-Gene; CADD: Scaled Combined Annotation Dependent Depletion v1.6; Impact: based on the MSC for the prioritized gene by V2G score. The MSC for *TAPBP*= 4.26, *C4orf33*= 4.87 and *COL11A2* =8.34 | | | | | | | | | |

| **Table S8. Significant eQTLs for *TAPBP* gene for the independent sentinel variants of 6p21.32.** | | | | |
| --- | --- | --- | --- | --- |
| Tissue | | rsID | p-value | NES* |
| Artery | Aorta | rs213226 | 1.70x10^-13^ | 0.29 |
|  |  | rs456261 | 9.50x10^-23^ | -0.38 |
|  |  | rs2076775 | 2.40x10^-9^ | 0.26 |
|  | Coronary | rs213226 | - | - |
|  |  | rs456261 | 1.90x10^-14^ | -0.38 |
|  |  | rs2076775 | 2.50x10^-6^ | 0.26 |
|  | Tibial | rs213226 | 3.30x10^-16^ | 0.22 |
|  |  | rs456261 | 1.70x10^-36^ | -0.33 |
|  |  | rs2076775 | 2.90x10^-10^ | 0.18 |
| Esophagus | Mucosa | rs213226 | 1.80x10^-9^ | 0.11 |
|  |  | rs456261 | 4.40x10^-12^ | -0.13 |
|  |  | rs2076775 | - | - |
|  | Muscularis | rs213226 | 5.40x10^-5^ | 0.12 |
|  |  | rs456261 | - | - |
|  |  | rs2076775 | - | - |
| Lung | | rs213226 | 3.30x10^-5^ | 0.092 |
|  |  | rs456261 | 1.50x10^-5^ | -0.096 |
|  |  | rs2076775 | - | - |
| Whole Blood | | rs213226 | - | - |
|  |  | rs456261 | 2.00x10^-7^ | 0.072 |
|  |  | rs2076775 | 4.00x10^-5^ | -0.06 |
| *Normalized effect size. Data obtained from: GTEx Analysis Release V8 (<https://gtexportal.org/>) | | | | |

| **Table S9. Top five most significant findings of the HLA associations for the amino acids and the classical alleles.** | | | | | | |
| --- | --- | --- | --- | --- | --- | --- |
| Type of variation | Gene | Variant | Position | OR[95% CI] | *p*-value | Freq. |
| Amino acids | *HLA-B* | 178 GK (exon 3) | 31323958 | 1.48[1.13-1.93] | 4.18x10^-3^ | 0.11 |
|  | *HLA-B* | 178 GT (exon 3) | 31323958 | 1.48[1.13-1.93] | 4.18x10^-3^ | 0.11 |
|  | *HLA-B* | 178 K (exon 3) | 31323958 | 1.48[1.13-1.93] | 4.18x10^-3^ | 0.11 |
|  | *HLA-B* | 178 T (exon 3) | 31323958 | 1.48[1.13-1.93] | 4.18x10^-3^ | 0.11 |
|  | *HLA-DPB1* | 65 L (exon 2) | 33048634 | 1.37[1.09-1.71] | 6.04x10^-3^ | 0.18 |
| Classical alleles | *HLA-A* | 31:01 | 29910607 | 1.90[1.15-3.13] | 0.012 | 0.02 |
|  | *HLA-A* | 32:01 | 29912530 | 0.48[0.25-0.92] | 0.027 | 0.04 |
|  | *HLA-B* | 07:02 | 31321650 | 1.40[1.02-1.91] | 0.037 | 0.07 |
|  | *HLA-B* | 40:01 | 31321949 | 1.77[1.08-2.89] | 0.023 | 0.02 |
|  | *HLA-DPB1* | 06:01 | 33043725 | 2.06[1.17-3.65] | 0.013 | 0.02 |
| Position: chromosome and base pair position according to GRCh37/hg19; OR: odd ratio; CI: confidence interval; Freq.: frequency. | | | | | | |

**Supplementary References:**

Calandra, T., & Cohen, J. (2005). The International Sepsis Forum Consensus Conference on Definitions of Infection in the Intensive Care Unit. Critical Care Medicine, 33(7), 1538–1548. https://doi.org/10.1097/01.CCM.0000168253.91200.83

Scicluna, B. P., Klein Klouwenberg, P. M. C., van Vught, L. A., Wiewel, M. A., Ong, D. S. Y., Zwinderman, A. H., Franitza, M., Toliat, M. R., Nürnberg, P., Hoogendijk, A. J., Horn, J., Cremer, O. L., Schultz, M. J., Bonten, M. J., & van der Poll, T. (2015). A Molecular Biomarker to Diagnose Community-acquired Pneumonia on Intensive Care Unit Admission. American Journal of Respiratory and Critical Care Medicine, 192(7), 826–835. https://doi.org/10.1164/rccm.201502-0355OC
