## Supplementary Table S6 for "A genome-wide association study of adults with community-acquired pneumonia"

**Table S6. Functional annotation of the variants with high biological impact included in the credible sets of the Bayesian fine mapping of 6p21.32**

| rsID | Position | Freq. (EUR) | Score [regulomeDB] | Rank [regulomeDB] | H3K4me1 and H3K27ac enhancer histone marks [HaploReg] | H3K4me3 and H3K9ac promoter histone marks [HaploReg] | DNAse [HaploReg] | Altered regulatory motifs [HaploReg] | Proteins bound [HaploReg] | eQTLs [HaploReg] | Score CAPE eQTL | CADD and MSC |
|---|---|---|---|---|---|---|---|---|---|---|---|---|
| rs456261 | chr6:33258443 | 0,524 | 0,554 | eQTL/caQTL + TF binding / chromatin accessibility peak (1f) | IMR90 fetal lung fibroblasts Cell Line, HUES6 Cells, HUES48 Cells, HUES64 Cells, ES-UCSF4 Cells, iPS-20b Cells, , hESC Derived CD56+ Mesoderm Cultured Cells, hESC Derived CD56+ Ectoderm Cultured Cells, hESC Derived CD184+ Endoderm Cultured Cells, Primary mononuclear cells from peripheral blood, Primary T cells from peripheral blood, Primary T cells effector/memory enriched from peripheral blood, Primary T cells from cord blood, Primary T regulatory cells from peripheral blood, Primary T helper cells from peripheral blood, Primary T helper naive cells from peripheral blood, Primary T helper cells PMA-I stimulated, Primary T helper 17 cells PMA-I stimulated, Primary T helper memory cells from peripheral blood 1, Primary T helper memory cells from peripheral blood 2, Primary T CD8+ memory cells from peripheral blood, Primary T CD8+ naive cells from peripheral blood, Primary monocytes from peripheral blood, Primary B cells from cord blood, Primary hematopoietic stem cells, Primary hematopoietic stem cells G-CSF-mobilized Male/Female, Primary hematopoietic stem cells short term culture, Primary B cells from peripheral blood, Primary Natural Killer cells from peripheral blood, Primary neutrophils from peripheral blood, Fetal Lung, Lung, A549 EtOH 0.02pct Lung Carcinoma Cell Line, GM12878 Lymphoblastoid Cells, K562 Leukemia Cells, Monocytes-CD14+ RO01746 Primary Cells, NHLF Lung Fibroblast Primary Cells... | IMR90 fetal lung fibroblasts Cell Line, ES-WA7 Cells, H9 Cells, ES-I3 Cells, HUES6 Cells, HUES48 Cells, HUES64 Cells, H1 Cells, ES-UCSF4 Cells, iPS-20b Cells, iPS-18 Cells, iPS-15b Cells, iPS DF 19.11 Cells, hESC Derived CD56+ Mesoderm Cultured Cells, hESC Derived CD56+ Ectoderm Cultured Cells, hESC Derived CD184+ Endoderm Cultured Cells, Primary mononuclear cells from peripheral blood, Primary T cells from peripheral blood, Primary T cells effector/memory enriched from peripheral blood, Primary T cells from cord blood, Primary T regulatory cells from peripheral blood, Primary T helper cells from peripheral blood, Primary T helper naive cells from peripheral blood, Primary T helper cells PMA-I stimulated, Primary T helper 17 cells PMA-I stimulated, Primary T helper memory cells from peripheral blood 1, Primary T helper memory cells from peripheral blood 2, Primary T CD8+ memory cells from peripheral blood, Primary B cells from cord blood, Primary hematopoietic stem cells G-CSF-mobilized Male/Female, Primary hematopoietic stem cells short term culture, Fetal Lung, A549 EtOH 0.02pct Lung Carcinoma Cell Line, GM12878 Lymphoblastoid Cells, Monocytes-CD14+ RO01746 Primary Cells, NHLF Lung Fibroblast Primary Cells... | None | None | None | DAXX, TAPBP, HLA-DPA1, WDR46, ZBTB22, B3GALT4, HLA-DPB1, SYNGAP1\|ZBTB9, VPS52 | 0.143 | 5.67 (MSC=4.26) |
| rs211476 | chr6:33319596 | 0,301 | 0,590 | TF binding or DNase peak (5) | None | None | None | NF-I_2, Smad3_2 | None | RPS18, TAPBP, DAXX, BAK1, HLA-DPA1, B3GALT4, CUTA, HSD17B8, SYNGAP1\|ZBTB9, VPS52 | 0.273 | 4.46 (MSC=4.26) |
| rs211475 | chr6:33320089 | 0,301 | 0,184 | Other (7) | None | None | None | Ik-1_2 | None | RPS18, TAPBP, DAXX, BAK1, HLA-DPA1 | 0.257 | 7.84 (MSC=4.26) |
| rs211473 | chr6:33320759 | 0,300 | 0,184 | Other (7) | None | None | None | RXRA_known7 | None | RPS18, DAXX, BAK1, HLA-DPA1 | 0.139 | 9.19 (MSC=4.26) |
| rs381847 | chr6:33401124 | 0,386 | 0,587 | TF binding + any motif + DNase Footprint + DNase peak (2b) | IMR90 fetal lung fibroblasts Cell Line, hESC Derived CD56+ Mesoderm Cultured Cells, hESC Derived CD56+ Ectoderm Cultured Cells, hESC Derived CD184+ Endoderm Cultured Cells, Primary mononuclear cells from peripheral blood, Primary T regulatory cells from peripheral blood, Primary T cells from peripheral blood, Primary T cells effector/memory enriched from peripheral blood, Primary T cells from cord blood, Primary T helper cells from peripheral blood, Primary T helper naive cells from peripheral blood, Primary T helper cells PMA-I stimulated, Primary T helper 17 cells PMA-I stimulated, Primary T helper memory cells from peripheral blood 2, Primary T CD8+ memory cells from peripheral blood, Primary T CD8+ naive cells from peripheral blood, Primary monocytes from peripheral blood, Primary B cells from cord blood, Primary hematopoietic stem cells, Primary hematopoietic stem cells G-CSF-mobilized (Male and Female), Primary hematopoietic stem cells short term culture, Primary B cells from peripheral blood, Primary Natural Killer cells from peripheral blood, Primary neutrophils from peripheral blood, Esophagus, Fetal Lung, Lung, Spleen, A549 EtOH 0.02pct Lung Carcinoma Cell Line, Dnd41 TCell Leukemia Cell Line, K562 Leukemia Cells, Lung, Monocytes-CD14+ RO01746 Primary Cells, NHLF Lung Fibroblast Primary Cells... | IMR90 fetal lung fibroblasts Cell Line, hESC Derived CD56+ Mesoderm Cultured Cells, hESC Derived CD56+ Ectoderm Cultured Cells, hESC Derived CD184+ Endoderm Cultured Cells, Primary T regulatory cells from peripheral blood, Primary T helper cells from peripheral blood, Primary T CD8+ naive cells from peripheral blood, Primary B cells from cord blood, Primary hematopoietic stem cells, Primary hematopoietic stem cells G-CSF-mobilized Male, Primary hematopoietic stem cells G-CSF-mobilized Female, Primary hematopoietic stem cells short term culture, Primary B cells from peripheral blood, Primary mononuclear cells from peripheral blood, Fetal Lung, GM12878 Lymphoblastoid Cells, Dnd41 TCell Leukemia Cell Line, Monocytes-CD14+ RO01746 Primary Cells ... | H1 BMP4 Derived Mesendoderm Cultured Cells, Primary B cells from peripheral blood, Fetal Thymus, Fetal Muscle Leg, Fetal Heart | None | POL2 | HLA-DPB2, TAPBP, DAXX, HLA-DPA1, HLA-DPB1 | 0.578 | 10.73 (MSC=4.26) |
| rs2247385 | chr6:33421577 | 0,386 | 0,705 | TF binding + DNase peak (4) | IMR90 fetal lung fibroblasts Cell Line, hESC Derived CD56+ Mesoderm Cultured Cells, hESC Derived CD56+ Ectoderm Cultured Cells, hESC Derived CD184+ Endoderm Cultured Cells, Primary mononuclear cells from peripheral blood, Primary T cells from peripheral blood, Primary T cells effector/memory enriched from peripheral blood, Primary T cells from cord blood, Primary T regulatory cells from peripheral blood, Primary T helper cells from peripheral blood, Primary T helper naive cells from peripheral blood, Primary T helper cells PMA-I stimulated, Primary T helper 17 cells PMA-I stimulated, Primary T helper memory cells from peripheral blood 1, Primary T helper memory cells from peripheral blood 2, Primary T CD8+ memory cells from peripheral blood, Primary T CD8+ naive cells from peripheral blood, Primary monocytes from peripheral blood, Primary B cells from cord blood, Primary hematopoietic stem cells, Primary hematopoietic stem cells G-CSF-mobilized (Male and Female), Primary hematopoietic stem cells short term culture, Primary B cells from peripheral blood, Primary Natural Killer cells from peripheral blood, Primary neutrophils from peripheral blood, Fetal Lung, Esophagus, Lung, Spleen, A549 EtOH 0.02pct Lung Carcinoma Cell Line, Dnd41 TCell Leukemia Cell Line, GM12878 Lymphoblastoid Cells, K562 Leukemia Cells, Monocytes-CD14+ RO01746 Primary Cells, NHLF Lung Fibroblast Primary Cells... | IMR90 fetal lung fibroblasts Cell Line, hESC Derived CD56+ Mesoderm Cultured Cells, hESC Derived CD56+ Ectoderm Cultured Cells, hESC Derived CD184+ Endoderm Cultured Cells, Primary mononuclear cells from peripheral blood, Primary T cells from peripheral blood, Primary T cells effector/memory enriched from peripheral blood, Primary T cells from cord blood, Primary T regulatory cells from peripheral blood, Primary T helper cells from peripheral blood, Primary T helper naive cells from peripheral blood, Primary T helper cells PMA-I stimulated, Primary T helper 17 cells PMA-I stimulated, Primary T helper memory cells from peripheral blood, Primary T CD8+ memory cells from peripheral blood, Primary T CD8+ naive cells from peripheral blood, Primary monocytes from peripheral blood, Primary B cells from cord blood, Primary hematopoietic stem cells, Primary hematopoietic stem cells G-CSF-mobilized (Male and Female), Primary hematopoietic stem cells short term culture, Primary B cells from peripheral blood, Primary Natural Killer cells from peripheral blood, Primary neutrophils from peripheral blood, Esophagus, Fetal Lung, Lung, Spleen, A549 EtOH 0.02pct Lung Carcinoma Cell Line, Dnd41 TCell Leukemia Cell Line, GM12878 Lymphoblastoid Cells, K562 Leukemia Cells, Monocytes-CD14+ RO01746 Primary Cells, NHLF Lung Fibroblast Primary Cells... | H9 Cells, Primary T cells from peripheral blood, Primary T cells from cord blood, Primary hematopoietic stem cells G-CSF-mobilized Male, Primary Natural Killer cells from peripheral blood, Foreskin Fibroblast Primary Cells skin02, Fetal Stomach, GM12878 Lymphoblastoid Cells, NH-A Astrocytes Primary Cells | ATF4, CTCF_disc5, DEC, E2F_disc1, Mxi1_known1, Rad21_disc7 | None | HLA-DPB2, TAPBP, DAXX, HLA-DPA1, HLA-DPB1, KIFC1, B3GALT4, CUTA, HSD17B8, ITPR3, PHF1, SYNGAP1\|ZBTB9 | 0.252 | 9.41 (MSC=4.26) |
| rs1755048 | chr6:33432160 | 0,386 | 0,135 | TF binding or DNase peak (5) | None | None | None | CEBPB_known2, Hic1_3, NF-I_1, Rad21_disc9, Zfp740 | None | HLA-DPB2, TAPBP, DAXX, HLA-DPA1, HLA-DPB1, B3GALT4, CUTA, ITPR3, PHF1, SYNGAP1\|ZBTB9 | 0.116 | 5.04 (MSC=4.26) |
| rs9283903 | chr6:33439777 | 0,394 | 0,342 | TF binding or DNase peak (5) | None | ES-WA7 Cells | None | YY1_known6 | None | TAPBP, DAXX, HLA-DPA1, HLA-DPB1, HLA-DPB2 | 0.128 | 5.45 (MSC=4.26) |
| rs2772393 | chr6:33443189 | 0,394 | 0,184 | Other (7) | H1 Derived Mesenchymal Stem Cells | Primary T CD8+ naive cells from peripheral blood, and iPS DF 19.11 Cells | None | DEC, Nkx2_7 | None | TAPBP, DAXX, HLA-DPA1, HLA-DPB1, HLA-DPB2 | 0.087 | 6.68 (MSC=4.26) |
| rs9366821 | chr6:33444074 | 0.385 | 0,004 | Motif hit (6) | None | H9 Cells, iPS DF 19.11 Cells, and Breast variant Human Mammary Epithelial Cells (vHMEC) | None | AP-2rep, MZF1::1-4_1 | None | TAPBP, DAXX, HLA-DPA1, HLA-DPB1, B3GALT4, CUTA, PHF1, SYNGAP1\|ZBTB9 | 0.126 | 14.62 (MSC=4.26) |

CADD: Combined Annotation Dependent Depletion;CAPE: CellulAr dePendent dEactivating mutations; caQTL: quantitative trait loci for chromatin accessibility; eQTL: Expression quantitative trait loci; hESC: human embryonic stem cells; iPS DF: induced pluripotent stem cell line derived from foreskin fibroblasts; MSC: Mutation Significance Cutoff; TF: transcription factor.
Position: chromosome and base pair according to GRCh37/hg19 build; Freq.: Frequency for non-Finish European, gnomAD genome data. The MSC for variants on chromosome 6 was estimated for the TAPBP gene prioritized with V2G (Variant-to-Gene).
