## Supplementary Table S7 for "A genome-wide association study of adults with community-acquired pneumonia"

**Table S7. Functional annotation of the variants with high biological impact included in the credible sets of the Bayesian fine mapping of 4q28.2**

| rsID | Position | Freq. (EUR) | Score [regulomeDB] | Rank [regulomeDB] | H3K4me1 and H3K27ac enhancer histone marks [HaploReg] | H3K4me3 and H3K9ac promoter histone marks [HaploReg] | DNAse [HaploReg] | Altered regulatory motifs [HaploReg] | Proteins bound [HaploReg] | eQTLs [HaploReg] | Score CAPE eQTL | CADD and MSC |
|---|---|---|---|---|---|---|---|---|---|---|---|---|
| rs34955650 | chr4:130254264 | 0,048 | 0,18 | Other (7) | H9 Derived Neuron Cultured Cells, H1 Derived Mesenchymal Stem Cells, Fetal Muscle Trunk | None | None | None | None | None | 0.608 | 7.75 (MSC=4.87) |
| rs17014611 | chr4:130255450 | 0,048 | 0 | TF binding or DNase peak (5) | HUES48 Cells, hESC Derived CD56+ Ectoderm Cultured Cells, hESC Derived CD184+ Endoderm Cultured Cells, H9 Derived Neuron Cultured Cells, Mesenchymal Stem Cell Derived Adipocyte Cultured Cells, Fetal Brain Female, Fetal Muscle Trunk, Fetal Muscle Leg, Fetal Kidney, Fetal Lung, and Pancreatic Islets. | ES-WA7 Cells | Fetal Muscle Trunk, Fetal Muscle Leg, Fetal Lung | Brachyury_2, Irf_known4 | None | None | 0.233 | 18.23 (MSC=4.87) |
| rs13109938 | chr4:130256289 | 0,048 | 0 | TF binding or DNase peak (5) | HUES48 Cells, ES-UCSF4 Cells, iPS-18 Cells, H1 Derived Neuronal Progenitor Cultured Cells, H9 Derived Neuronal Progenitor Cultured Cells, H9 Derived Neuron Cultured Cells, hESC Derived CD56+ Ectoderm Cultured Cells, hESC Derived CD184+ Endoderm Cultured Cells, Mesenchymal Stem Cell Derived Adipocyte Cultured Cells, Ganglion Eminence derived primary cultured neurospheres, Fetal Kidney, Fetal Lung, Pancreatic Islets... | HUES48 Cells, HUES64 Cells, Adipose Nuclei, ES-WA7 Cells, Pancreatic Islets | None | Cdx, Dbx1, Fox, Foxa_known1, Foxa_known2, Foxa_known4, Foxd3, Foxf1, Foxi1, Foxj1_1, Foxj1_2, Foxj2_1, Foxl1_1, Foxo_3, HDAC2_disc2, HDAC2_disc6, Hlx1, Mef2_disc1, Nkx6-2, Pax-6_1, Pou3f2_2, STAT_known11, p300_disc5 | None | None | 0.182 | 19.34 (MSC=4.87) |
| rs35004602 | chr4:130256929 | 0,048 | 0,81 | TF binding + any motif + DNase Footprint + DNase peak (2b) | ES-I3 Cells, HUES48 Cells, HUES64 Cells, ES-UCSF4 Cells, iPS-20b Cells, iPS-18 Cells, iPS-15b Cells, H1 Derived Neuronal Progenitor Cultured Cells, H9 Derived Neuronal Progenitor Cultured Cells, H9 Derived Neuron Cultured Cells, hESC Derived CD56+ Ectoderm Cultured Cells, hESC Derived CD184+ Endoderm Cultured Cells, Mesenchymal Stem Cell Derived Adipocyte Cultured Cells, Fetal Kidney, Fetal Lung, Pancreatic Islets, HSMM cell derived Skeletal Muscle Myotubes Cells... | HUES48 Cells, HUES64 Cells, iPS-20b Cells, Adipose Nuclei, and Brain Anterior Caudate | Fetal Muscle Leg, Fetal Lung | HNF4_known1, HNF4_known4, PPAR_3, PTF1-beta, RREB-1_1, RXRA_known1, RXRA_known6 | None | None | 0.206 | 15.70 (MSC=4.87) |
| rs17014614 | chr4:130269324 | 0,047 | 0,18 | Other (7) | Foreskin Melanocyte Primary Cells skin03, Brain_Dorsolateral_Prefrontal_Cortex | Breast Myoepithelial Primary Cells | None | None | None | None | 0,131 | 11.03 (MSC=4.87) |
| rs13128188 | chr4:130276663 | 0,048 | 0,61 | TF binding + chromatin accessibility peak (4) | ES-I3 Cells, HUES6 Cells, HUES48 Cells, HUES64 Cells, ES-UCSF4 Cells, iPS-20b Cells, iPS-15b Cells, iPS DF 6.9 Cells, H9 Derived Neuronal Progenitor Cultured Cells, H9 Derived Neuron Cultured Cells, hESC Derived CD56+ Mesoderm Cultured Cells, hESC Derived CD56+ Ectoderm Cultured Cells, hESC Derived CD184+ Endoderm Cultured Cells, Fetal Lung... | Adipose Derived Mesenchymal Stem Cell Cultured Cells, Brain Anterior Caudate, Fetal Heart | Fetal Brain Female, Fetal Brain Male, Fetal Muscle Leg, Fetal Heart | Pax-8_1, Pou2f2_disc1 | None | None | 0,753 | 11.87 (MSC=4.87) |
| rs35524133 | chr4:130277071 | 0,047 | 0,61 | TF binding + chromatin accessibility peak (4) | ES-I3 Cells, HUES6 Cells, HUES48 Cells, HUES64 Cells, ES-UCSF4 Cells, iPS-20b Cells, iPS-15b Cells, iPS DF 6.9 Cells, H9 Derived Neuronal Progenitor Cultured Cells, H9 Derived Neuron Cultured Cells, hESC Derived CD56+ Mesoderm Cultured Cells, hESC Derived CD56+ Ectoderm Cultured Cells, hESC Derived CD184+ Endoderm Cultured Cells, Fetal Lung... | Adipose Derived Mesenchymal Stem Cell Cultured Cells, Brain Anterior Caudate, Fetal Heart | Fetal Heart | None | None | None | 0,19 | 9.10 (MSC=4.87) |
| rs28580176 | chr4:130281632 | 0,054 | 0,13 | TF binding or chromatin accessibility peak (5) | HUES48 Cells, Primary hematopoietic stem cells short term culture, Adipose Derived Mesenchymal Stem Cell Cultured Cells, Mesenchymal Stem Cell Derived Adipocyte Cultured Cells, Fetal Lung, HSMM cell derived Skeletal Muscle Myotubes Cells, Pancreas | Primary mononuclear cells from peripheral blood, Primary T CD8+ naive cells from peripheral blood, Brain Anterior Caudate | None | BATF_disc2, Irf_disc6, Irf_known6, TCF4_disc2, TCF4_known2, TCF4_known3 | None | None | 0,261 | 8.26 (MSC=4.87) |
| rs11942914 | chr4:130288620 | 0,053 | 0,59 | Motif hit (6) | Stomach Smooth Muscle, Pancreatic Islets | None | None | Dbx1, HNF1_1, HNF1_6, Hoxa9, Hoxc6, Ncx_2, Nkx6-1_1, Nkx6-1_2, Pax-6_3, Pou4f3 | None | ENSG00000151470.7_130023757_130023946 | 0,24 | 5.49 (MSC=4.87) |
| rs13152421 | chr4:130291896 | 0,053 | 0,13 | TF binding or DNase peak (5) | hESC Derived CD184+ Endoderm Cultured Cells, H1 Derived Neuronal Progenitor Cultured Cells, H9 Derived Neuronal Progenitor Cultured Cells, H9 Derived Neuron Cultured Cells, Fetal Heart, Duodenum Smooth Muscle, Colon Smooth Muscle, Rectal Smooth Muscle, Fetal Stomach, Pancreatic Islets, Brain Inferior Temporal Lobe, Fetal Muscle Leg | None | Fetal Muscle Leg | None | None | ENSG00000151470.7_130023757_130023946 | 0.301 | 14.26 (MSC=4.87) |
| rs35618244 | chr4:130298386 | 0,052 | 0,83 | TF binding + any motif + Footprint + chromatin accessibility peak (2b) | ES-WA7 Cells, hESC Derived CD56+ Mesoderm Cultured Cells, Primary hematopoietic stem cells G-CSF-mobilized Female, Mesenchymal Stem Cell Derived Chondrocyte Cultured Cells, Fetal Lung, Dnd41 TCell Leukemia Cell Line, HUVEC Umbilical Vein Endothelial Primary Cells... | Fetal Lung, Foreskin Keratinocyte Primary Cells skin02, Foreskin Keratinocyte Primary Cells skin03, NHEK-Epidermal Keratinocyte Primary Cells | Foreskin Keratinocyte Primary Cells skin02, Fetal Brain Female, HUVEC Umbilical Vein Endothelial Primary Cells, NHEK-Epidermal Keratinocyte Primary Cells | CIZ, FXR_1, Foxa_known2, Foxd3, Foxo_3, Foxp1, GATA_known8, HDAC2_disc6, Irf_disc3, Nanog_disc2, Sox_13, Sox_2, Sox_6, Sox_7, Zfp105, p300_disc5 | CFOS | None | 0.194 | 6.06 (MSC=4.87) |
| rs36062368 | chr4:130307035 | 0,053 | 0.18412 | Other (7) | Breast Myoepithelial Primary Cells, HMEC Mammary Epithelial Primary Cells, NHEK-Epidermal Keratinocyte Primary Cells | None | None | Irf_known2, Maf_known1, Mrg1::Hoxa9_1, Myf_4, TAL1_known2, TCF11::MafG | None | ENSG00000151470.7_130023757_130023946 | 0.16 | 5.78 (MSC=4.87) |
| rs13136673 | chr4:130360060 | 0,132 | 0,61 | TF binding + chromatin accessibility peak (4) | None | Liver | Fetal Lung, Fetal Adrenal Gland | AP-4_2, AP-4_3, Ascl2, E2A_2, E2A_5, LBP-1_2, Pou2f2_disc1, Pou3f2_1 | GATA2 | C4orf33 | 0.103 | 7.97 (MSC=4.87) |

CADD: Combined Annotation Dependent Depletion; CAPE: CellulAr dePendent dEactivating mutations; caQTL: quantitative trait loci for chromatin accessibility; eQTL: Expression quantitative trait loci; hESC: human embryonic stem cells; iPS DF: induced pluripotent stem cell line derived from foreskin fibroblasts; MSC: Mutation Significance Cutoff; TF: transcription factor.
Position: chromosome and base pair according to GRCh37/hg19 build; Freq.: Frequency for non-Finish European, gnomAD genome data. The MSC for variants on chromosome 6 was estimated for the TAPBP gene prioritized with V2G (Variant-to-Gene).
